# Integrating 16S and 18S sequencing reveals coupled prokaryotic and eukaryotic relationships in drinking water biofilms

**DOI:** 10.64898/2026.10.29.26366860

**Authors:** Laura Caligaris, Frederik Hammes, Sandra Cristino, Marco Gabrielli

## Abstract

Drinking water biofilms host diverse communities spanning all realms of life. However, despite the known interactions between opportunistic pathogens and microbial eukaryotes such as amoebae, most studies focus only on the prokaryotic populations in these systems. As a result, the degree to which microbial eukaryotes shape biofilm communities and the persistence of opportunistic waterborne pathogens, such as *Legionella* spp. is still largely unknown. Here we leveraged a set of 128 non-disinfected drinking water shower hose biofilms collected across Switzerland to characterize both communities and quantify eukaryotes’ role in shaping prokaryotic communities and *Legionella* persistence. We found both communities to present a conserved set of core taxa dominated by oligotrophic bacterial families and amoebae. The prevalence of taxa was mainly driven by the phylogenetic diversity among their sequence variants rather than their number alone. We detected prokaryotic and eukaryotic communities to show similar patterns in their richness and composition, influencing each other more than abiotic building-level factors. *Legionellaceae* bacteria were detected in 82% of samples presenting a diversity extending known reference species and no clear dominant taxa. *Legionellaceae* richness did not correlate with the richness of known amoebal hosts, but rather other amoebae, indicating that eukaryotic taxa relevant for *Legionella* persistence extend beyond the classically described hosts. These findings highlight the importance of integrating the eukaryotic communities to understand the microbial ecology in drinking water biofilms and recognize the conditions which allow opportunistic waterborne pathogens such as *Legionella* to persist.

## Introduction

Biofilms are structured communities of microorganisms embedded in a self-produced matrix of extracellular polymeric substances which favour strong cell-to-cell interactions. This environment also offers protection from environmental stresses, such as desiccation and disinfection, and retains nutrients from the water phase (Flemming & Wingender, 2010). In drinking water systems, biofilms can grow inside pipelines and outlets, in which prokaryotic and eukaryotic communities can establish (S. Liu et al., 2016).

Cooperation and competition shape the drinking water biofilm communities. Among the interactions between prokaryotes and eukaryotes, grazing by protists is a strong selective pressure on the bacterial community: many are consumed and digested, while others, called amoebae-resistant bacteria (ARB), survive and replicate inside their host, finding in this interaction an additional protection (Fan et al., 2024). In some instances this latter interaction also protects the eukaryotes, for example in amoebae, where some symbionts limit the invasion from other parasites (L. König et al., 2019). Across the ARB, waterborne pathogens such as *Legionella,* Nontuberculous Mycobacteria (NTM) and *Pseudomonas aeruginosa* can be found (Greub & Raoult, 2004). The amoebal host can protect them from biocides and other stresses, favouring their persistence and dispersion in drinking water distribution systems (DWDSs). This increases the risk exposure for the user and the healthcare-associated infections related to water (Cateau et al., 2014).

Most of the drinking water biofilm studies focus solely on prokaryotic (i.e., bacterial and archeal) diversity under different conditions such as temperature, disinfection treatment and seasonality (Healy et al., 2024; Mi et al., 2015; Moreno et al., 2024), while eukaryotic communities remain very poorly characterized, despite their role in the ecology of opportunistic pathogens. Eukaryotic diversity in DWDSs is shaped by both management and climatic factors and highly related to prokaryotic diversity, suggesting a close link between the two kingdoms (Gabrielli et al., 2023). The composition of eukaryotic communities is substantially different between water and biofilms, even though presenting in both cases a large fraction of unclassified sequences. Biofilms from different locations were different but functionally similar, representing also a hotspot for pathogenic amoebae (Li et al., 2021). Biofilms are, in particular, hotspots of diversity for eukaryotic species known to host ARBs (Inkinen et al., 2019). It is then critical to improve our understanding of the role of eukaryotes in DWDSs to improve current microbiological management strategies and protect public health (Gabrielli et al., 2023).

Despite different studies on drinking water microbiota, prokaryotic and eukaryotic communities together are rarely characterized in the same samples, and the taxa considered as relevant hosts for waterborne pathogens are largely limited to a small number of amoebae described in literature (Boamah et al., 2017). Hence, a combined characterization of both kingdoms in the same biofilms is required to understand the ecological context in which opportunist pathogens persist, particularly in non-disinfected water systems, which remain less studied than disinfected ones (Sudarshan et al., 2024).

Here we sequenced the prokaryotic and the eukaryotic communities of 128 non-disinfected drinking water shower hose biofilms collected across Switzerland, using full-length 16S rRNA and long-amplicon 18S-ITS-28S rRNA sequencing. We characterized the two communities separately, assessed their associations through diversity and co-occurrence network analyses. We also focused on *Legionella* and its relationships with the eukaryotic community, comparing the amoebae described as hosts in the literature with the taxa identified through network analysis.

## Results

### Non-disinfected drinking water biofilms host diverse bacteria and amoebae, with taxon prevalence driven by phylogenetic diversity

The shower hose biofilm samples revealed diverse prokaryotic and eukaryotic communities with several taxa present at high relative abundance across samples (Fig. 1, Fig. 2). We identified 15106 ASVs distributed across 472 families, with single samples harbouring between 60 and 1139 ASVs. Among all the detected ASVs, 44 ASVs belonging to 22 families presented high relative abundance (median = 0.52%) and prevalence (median = 59.9%) and constituted the core biofilm community (Fig. 1a) (Shade & Stopnisek, 2019). In particular, ASVs belonging to the families *Sphingomonadaceae*, *Burkholderiaceae*, *Xanthobacteraceae*, *Caulobacteraceae,* and *Pyrinomonadaceae* were found in each sample and presented the highest relative abundance. We also identified ASVs belonging to families that include waterborne opportunistic pathogens such as *Legionellaceae*, *Mycobacteriaceae* and *Pseudomonadaceae* (Fig. 1a). Among these, *Legionellaceae* was detected in 82% of samples, while *Mycobacteriaceae* (39%) and *Pseudomonadaceae* (35%) were less prevalent. The median of relative abundances was similar between *Legionellaceae* (0.071%) and *Pseudomonadaceae* (0.074%), while for *Mycobacteriaceae* was lower (0.030%). Despite the relatively large family-level prevalence, no ASV from these families were part of the core community at ASV-level indicating that the prevalence of these families is not driven by a single ASV.

**Figure 1:**
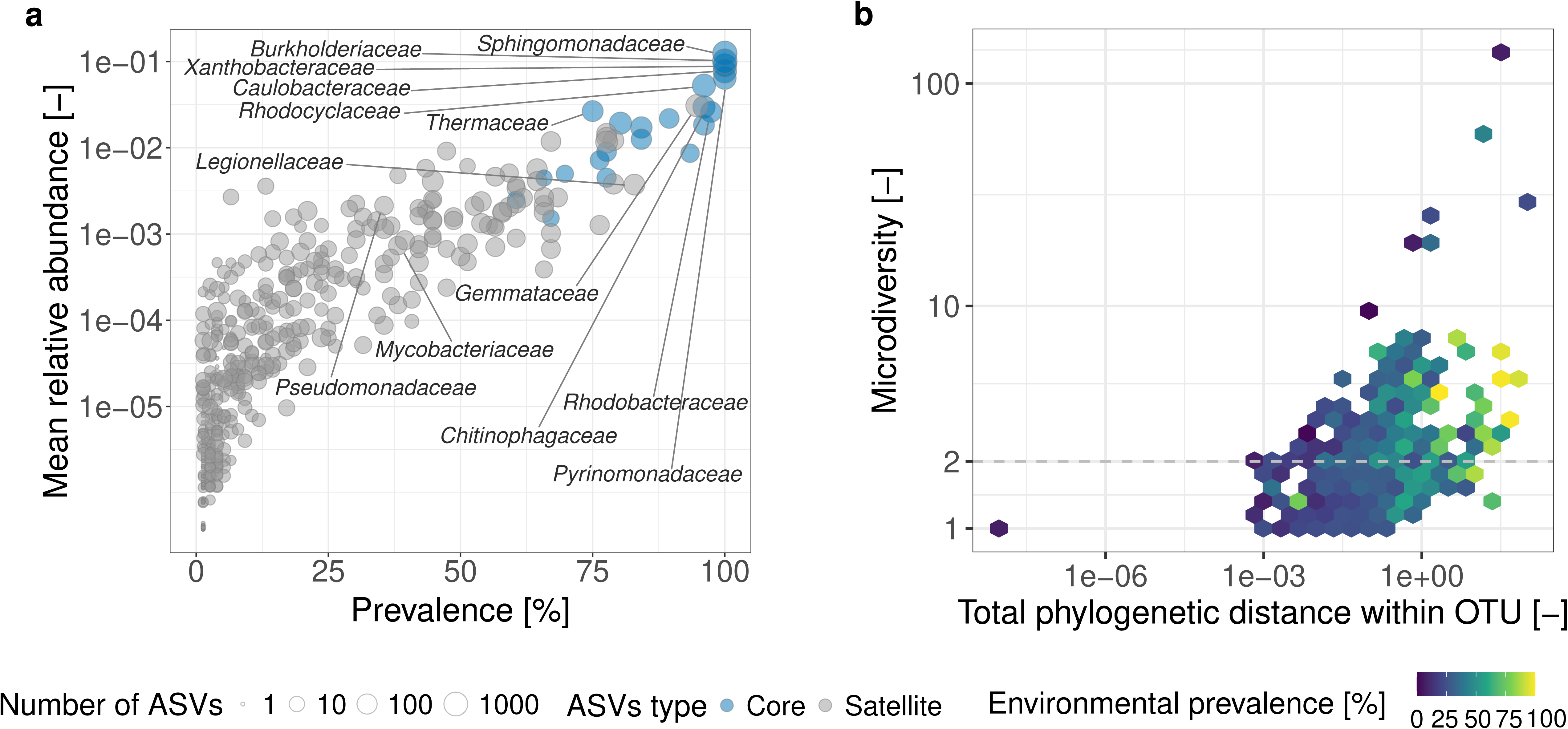
Prevalence and mean relative abundance of prokaryotic ASVs (a) and the relationship between prevalence, microdiversity and phylogenetic distance (b). (a) Each circle represents a family-level taxon and the point size indicates the number of ASVs per family. Families with at least one ASV in the core community are shown in blue, while families with all ASVs belonging to the satellite community are depicted in grey. The top 10 families by mean relative abundance are family-level labelled, while the families containing opportunistic pathogens, i.e. *Legionellaceae*, *Pseudomonadaceae* and *Mycobacteriaceae* are labelled regardless of abundance rank. (b) Microdiversity represents the effective number of ASVs within each OTU, while total phylogenetic distance is the sum of pairwise phylogenetic distances between the ASVs in each OTU. Color indicates the environmental prevalence of each OTU across samples. The dashed line indicates the microdiversity threshold of 2 effective ASVs.

**Figure 2:**
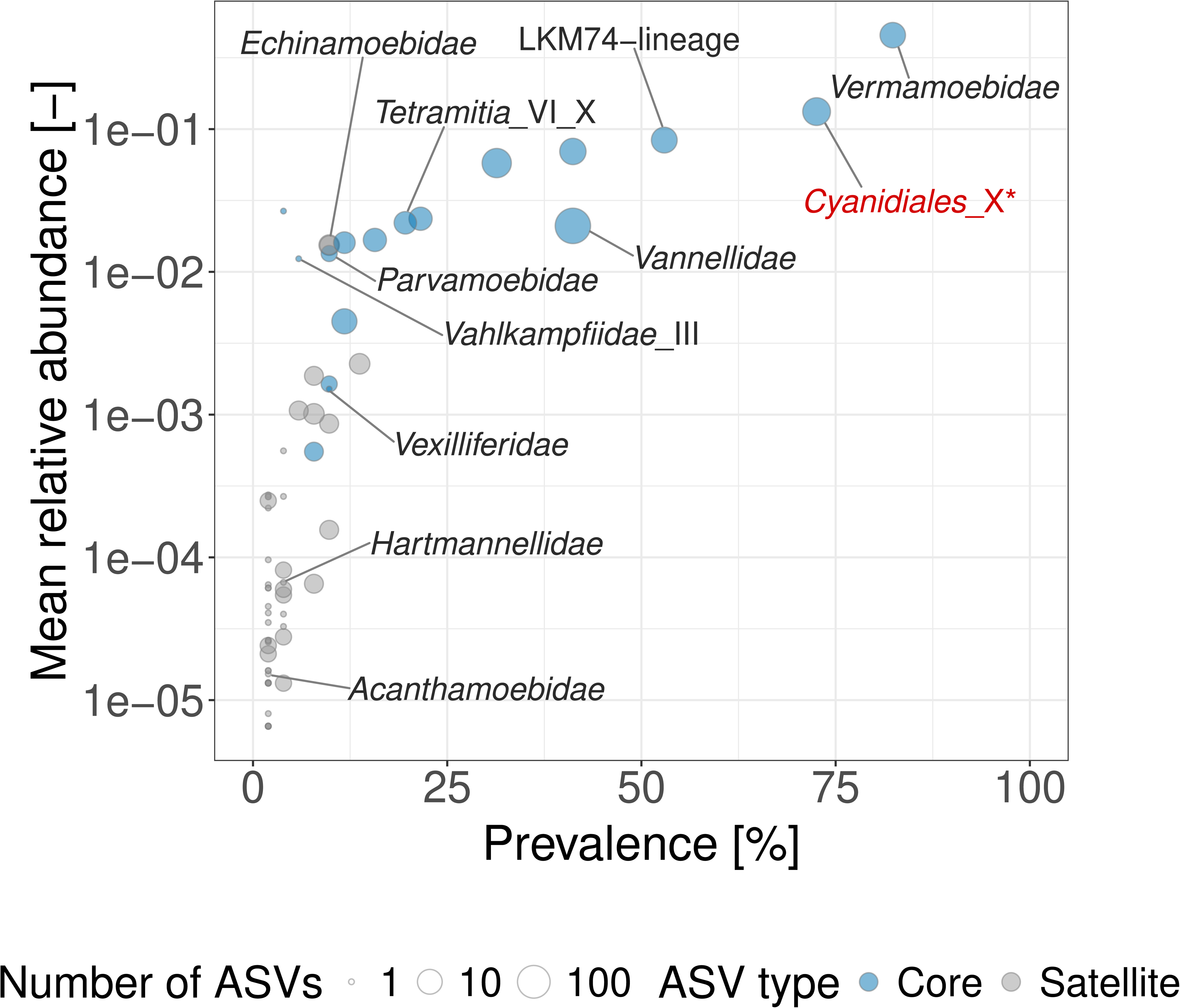
Prevalence and mean relative abundance of eukaryotic ASVs. Each circle represents a family-level taxon and point size is the number of ASVs per family. Families with at least one ASV in the core community in blue, while families with all ASVs belonging to the satellite community are shown in grey. Amoebae known as pathogen reservoirs are labelled. Asterisk and the red color indicate a presumed misclassification of *Cyanidiales*.

Across *Legionellaceae* ASVs (n = 154), only 44 received a species-level assignment and only 8 showed similarity greater than 98.7% to the classified ones (Fig. S2). To provide more information regarding the unclassified ASVs, we then reconstructed a phylogenetic tree of *Legionella* ASVs including 65 *Legionella* reference genomes (Fig. S7). Among the unclassified ASVs, 36 ASVs were placed within clades containing reference genomes, while 66 ASVs formed clades without any reference genome, highlighting the current lack of knowledge regarding environmental *Legionellae*. *L. waltersii* was the most prevalent species, detected in 45 of the samples where *Legionella* was identified (59% of all samples), and included 21 ASVs assigned to the species level, as well as 3 unassigned, but highly-related (similarity > 98.7%) ASVs. In concordance with the core community analysis, its presence was not sustained by a single dominant variant: the most widespread ASV was detected in 22 samples (28%), while 13 of the 24 ASVs were detected in only a single sample (Fig. S2). Besides *L. waltersii*, the most prevalent *Legionella* OTUs (i.e., >20%) were taxonomically unannotated and belonged to distinct clades compared to reference genomes, once again stressing our lack of understanding of the diversity of highly-prevalent environmental *Legionella* bacteria. On the contrary, *L. pneumophila* was detected in 4 samples (5%) with only 3 ASVs (Fig. S2), suggesting a surprisingly low prevalence of this clinically-relevant species despite being responsible for most of the cases in Switzerland (Fischer, Mäusezahl, et al., 2023).

To better understand the main drivers of the taxa distribution, we clustered ASVs into species-level operational taxonomic units (OTUs) using a similarity cutoff of 98.7% (Yarza et al., 2014). Then, we rarefied each OTU to a comparable number of reads, and calculated the OTU-level phylogenetic diversity of the retained ASVs. Finally, we estimate the Shannon diversity of each OTU and, similarly to previous work (García-García et al., 2019), defined this as its microdiversity (Fig. 1b). Of 631 OTUs for which we were able to estimate microdiversity, 229 had more than 2 effective ASVs and, in accordance to what previously reported (García-García et al., 2019), presented a higher environmental prevalence across samples (median = 35%) than those with microdiversity lower than 2 (median = 21%) (Wilcoxon test, *p* < 0.001). Besides microdiversity, we noticed how OTUs with higher phylogenetic distance (i.e., a measure of how phylogenetically divergent ASVs within the same OTU are) showed the highest environmental prevalence (Fig. 1b). We then used a Random Forest analysis to measure the relative contribution of microdiversity and phylogenetic distance on OTU prevalence. Our results show the phylogenetic distance within an OTU to provide about 4.5 times more information regarding OTU prevalence than microdiversity (Fig. S1b), suggesting that, despite the known limitations of 16S rRNA marker genes (e.g., Jaspers & Overmann, 2004), phylogenetically diverse OTUs are able to establish in more different communities (Malfertheiner et al., 2026).

We detected 627 eukaryotic ASVs, with 1 to 171 ASVs per sample (median = 15), distributed across 67 families. Based on ASVs prevalence and relative abundance, we identified 94 ASVs to form the core eukaryotic community (Fig. 2) and showing a median prevalence of 7.8% and relative abundance of 0.18%. Among these core ASVs, 61 belonged to 17 families, while 33 could not be assigned at family level. Most of the core ASVs belonged to amoebae generally known or proposed as reservoirs for pathogens, such as *Vermamoebidae* (82% prevalence, 45.5% mean relative abundance) and *Vannellidae* (41%, 2.1%), as well as LKM74-lineage (53%, 8.4%). The family *Vannellidae* showed the highest number of ASVs (n = 275), despite being detected in fewer samples. The only amoebae whose ASVs did not belong to the core community were *Acanthamoebidae*, *Hartmannellidae* and *Echinamoebidae*. Notably, 22 ASVs were erroneously classified as Cyanidiales_X (i.e., extremophilic algae; Stadnichuk & Tropin, 2022), a family with relatively high prevalence (72%) and median relative abundance (13.3%). In-depth analyses of these ASVs identified them as most similar to *Angulamoeba* sp. FN806 (Accession Number KP864086.1) (Table S1) and well-supported within its genus (Fig. S5), further reinforcing the picture of an amoeba-dominated eukaryotic community.

As performed for the prokaryotic dataset, we clustered the eukaryotic ASVs into OTU to estimate the relative contribution of microdiversity and phylogenetic diversity to their environmental prevalence. However, the limited sequencing depth of the eukaryotic dataset, allowed us to retain only a very limited number of OTUs (i.e., 34) precluding further analyses.

### Prokaryotic and eukaryotic communities show associated richness and composition across biofilms

Given the growing recognition regarding protist-prokaryote interactions (Cavallaro et al., 2023; Gabrielli et al., 2023), we then assessed whether the two communities showed similar patterns of richness and diversity across the sampled biofilms. The observed richness of the two communities (i.e., the total number of ASVs detected regardless of their abundance) showed a low but significant correlation (rho = 0.36, *p* = 0.02, Fig. 3a). The correlation between the two communities significantly increased when considering the Shannon diversity index (rho = 0.50, *p* < 0.001, Fig. 3b). Since this richness measure accounts for the evenness of taxa distribution, this result suggests that the two communities are coupled through their most abundant members.

**Figure 3:**
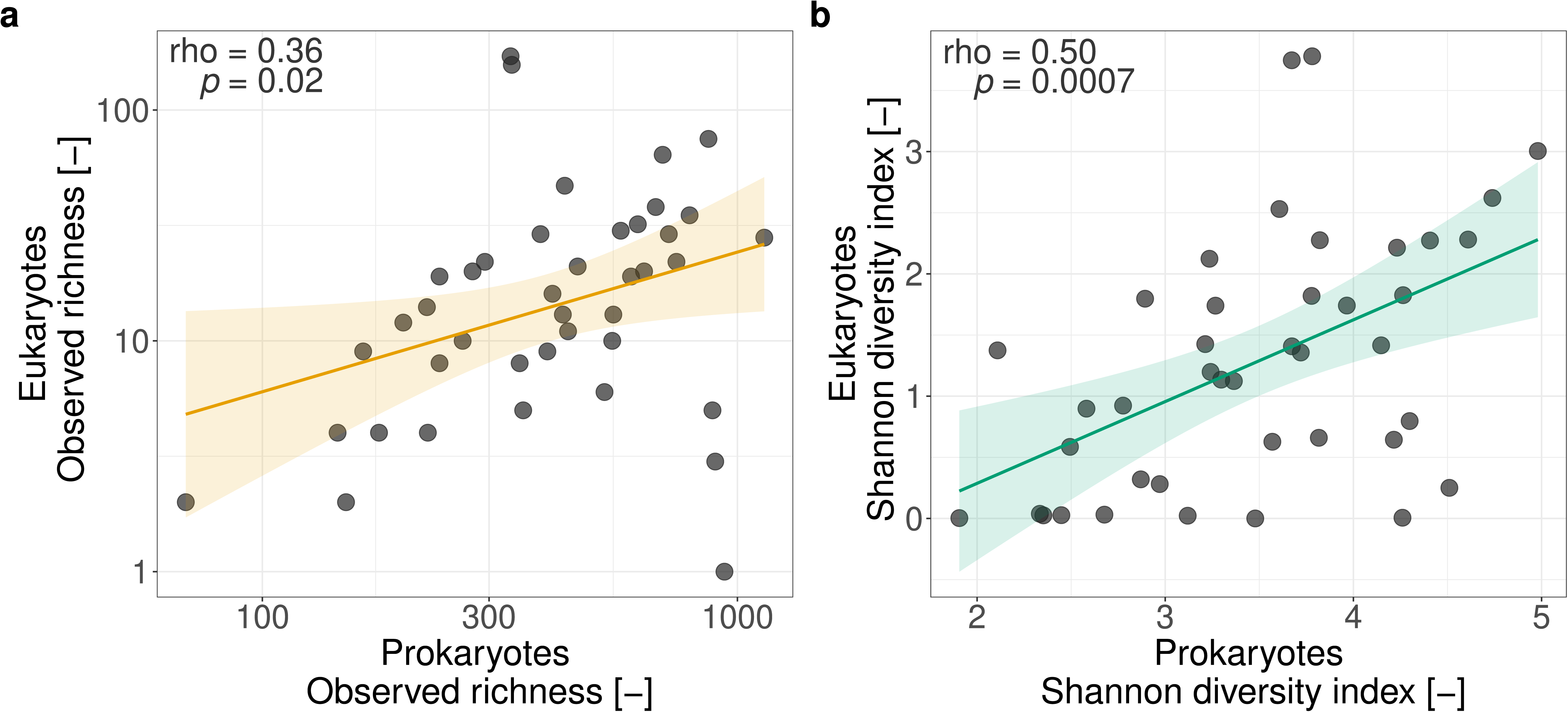
Observed richness (a) and Shannon diversity index (b) calculated for eukaryotes and prokaryotes. Each data point represents a sample shared between the two datasets. Coloured lines show linear fit between the two communities with yellow representing the observed richness (number of ASVs detected) and green the Shannon diversity index; shaded areas indicate the 95% confidence interval. Spearman correlation coefficients and p-values are shown in each panel.

We then moved to consider the composition of the biofilm communities, rather than only their richness. We performed a Procrustes analysis based on the principal component ordinations (PCoA) of Bray-Curtis community dissimilarities and identified significant congruence between the two ordinations (r = 0.318, *p* = 0.028, Fig. 4a), indicating that samples with similar prokaryotic composition tended to show similar eukaryotic composition. Outliers in the ordination corresponded to samples with very low eukaryotic richness (1-19 ASVs) despite normal prokaryotic richness (163-939 ASVs). Focusing on all members of both communities irrespective of their relative abundance using Jaccard dissimilarities resulted to a lower and non statistically significant linkage between the composition of the two communities (r = 0.219, *p* = 0.268, Table S2) highlighting, once again, how this phenomenon is driven by the most abundant taxa.

**Figure 4:**
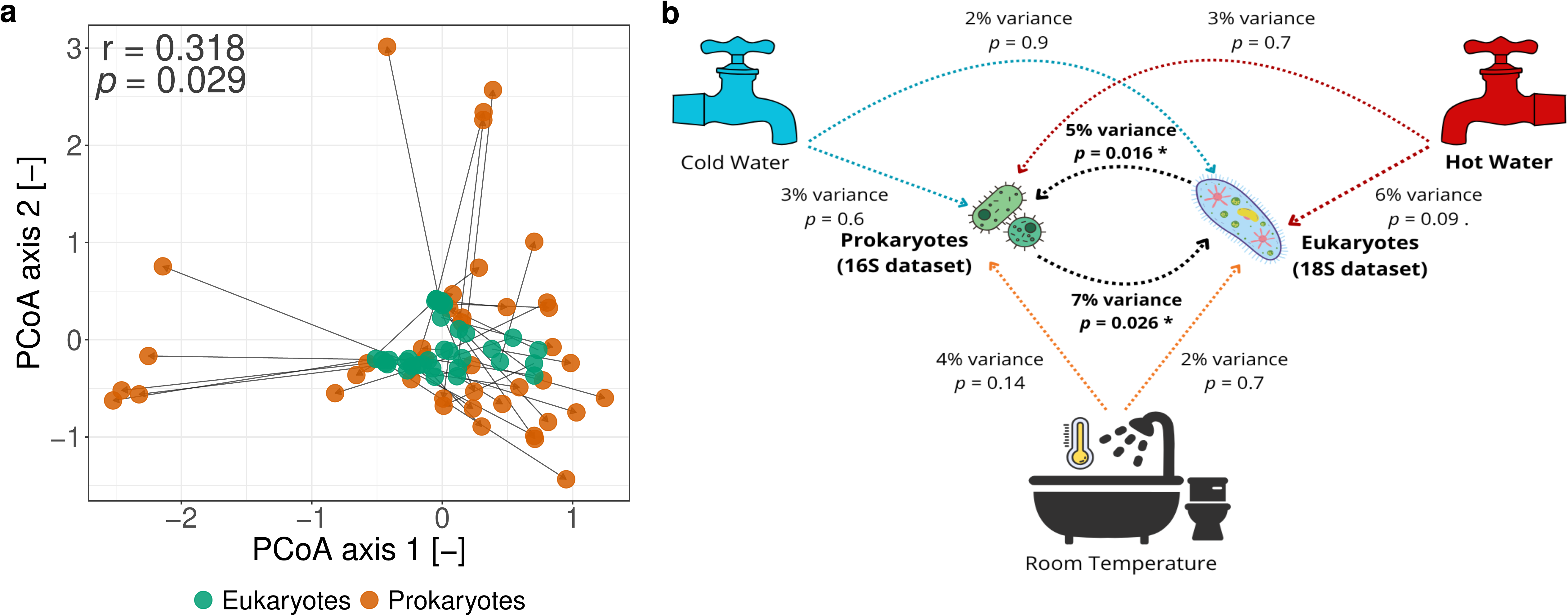
Beta diversity analysis between Prokaryotes and Eukaryotes communities (a) and influence of abiotic and biotic factors. (a) Procrustes analysis comparing the beta diversity structure of the Prokaryotes (orange) and Eukaryotes (green) communities based on Bray-Curtis dissimilarity. Each arrow connects the same sample in the two ordination spaces after optimal rotation. Shorter arrows indicate higher congruence between bacterial and eukaryotic community structures. Configurations were scaled prior to Procrustes rotation to account for differences in variance between the two datasets (r = 0.318, *p* = 0.029, permutations = 9999). (b) Influence of abiotic (cold water, hot water and room temperature) and biotic factors on prokaryotic and eukaryotic communities estimated using Permutational Multivariate Analysis of Variance (Bray-Curtis dissimilarity). Arrows indicate the direction of the tested effect. Bold values indicate statistically significant results (*p* < 0.05, *). Non-significant associations are shown in regular font.

We then used PERMANOVA tests to better quantify the linkage between the two communities and compare it to the effect of building-level factors. The community composition of both communities (measured as the first PCoA axis) showed a significant influence on the other: prokaryotes explained the 7% of variance in the the eukaryotic communities (*p* = 0.026), while, vice versa, eukaryotes explained 5% of variance of prokaryote ones (*p* = 0.016, Fig. 5b). Conversely, we found building factors (hot/cold water, room temperature) to have no significant explanatory power on neither community (Table S3), suggesting a much weaker effect compared to biotic interactions. Beyond building-level factors, we did not find a significant influence of water source on the community composition of neither kingdom, both when including municipalities fed with mixed water sources (PERMANOVA: prokaryotes R^2^ = 0.049, *p* = 0.276, eukaryotes R^2^ = 0.917, *p* = 0.577) as well as focusing only on groundwater and springwater dominated samples (prokaryotes R^2^ = 0.057, *p* = 0.097, eukaryotes R^2^ = 0.0594, *p* = 0.483).

**Figure 5:**
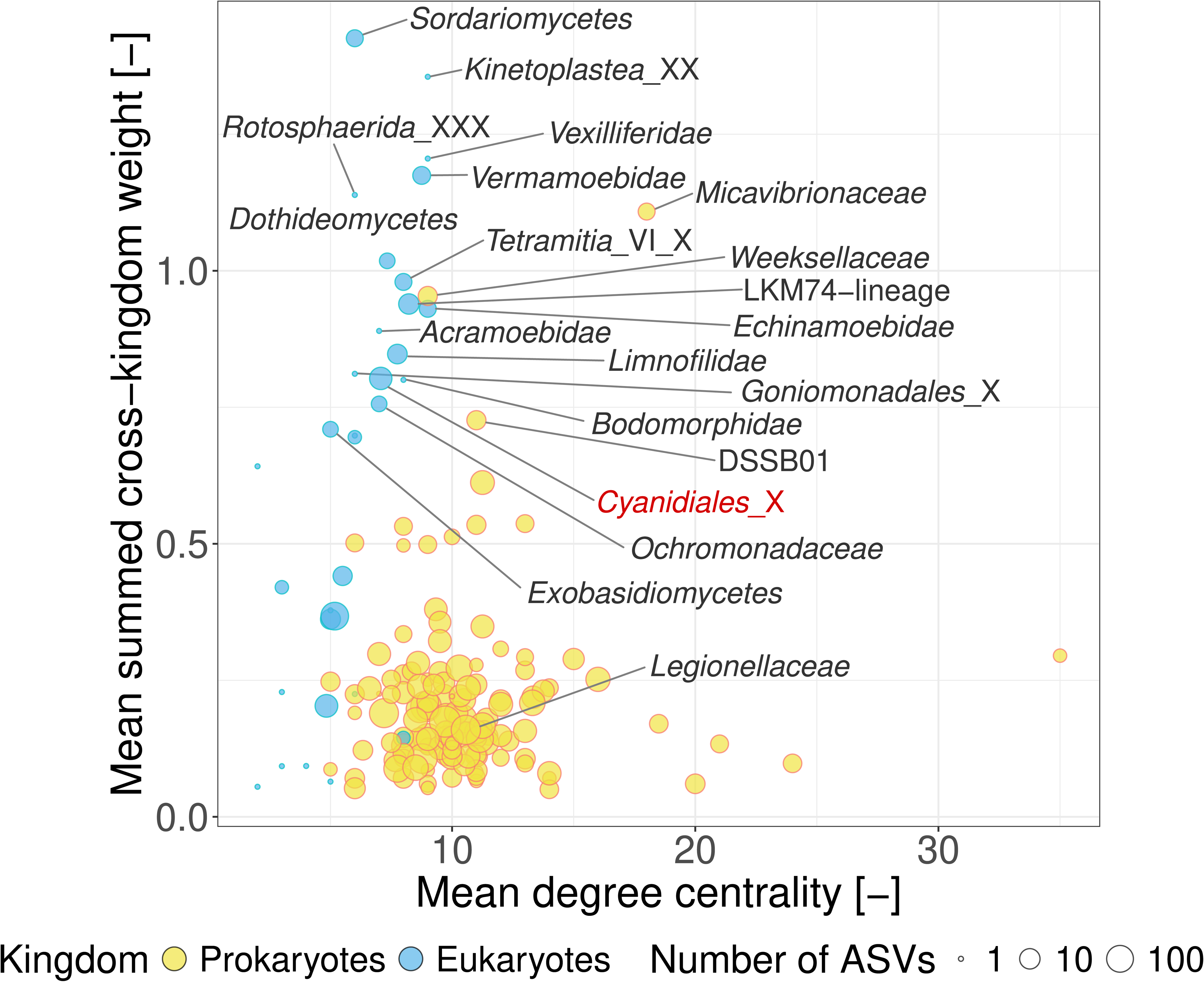
Cross-kingdom degree centrality and association weight for prokaryotic and eukaryotic families. Each point represents a family from the prokaryotes (in yellow) and the eukaryotes (in blue); the size of the point is the number of ASVs assigned to each family. On the x axis, the mean degree centrality is the number of connections each ASV has per family in the co-occurrence network. On the y axis, the mean summed cross-kingdom weight is the average sum of the network edge weights connecting each taxa to another from the other kingdom. Family labels are shown for mean summed cross-kingdom weights up to 0.70 and Legionellaceae. The higher cross-kingdom weight observed for the eukaryotic dataset is partially influenced by the larger number of nodes derived from the prokaryotic dataset in the network.

To understand which prokaryotic and eukaryotic taxa were the most involved in cross-kingdom interactions, we constructed a co-occurrence network and estimated the degree of centrality of each prokaryotic and eukaryotic family and the sum of the edge weights connecting it to the other kingdom (i.e., cross-kingdom weights; Fig. 5). Eukaryotic families (n = 35) showed higher mean summed cross-kingdom weights and lower mean degree of centrality than prokaryotic ones (n = 142). Among the them, the highest cross-kingdom weights were found for amoebae, including *Vexillifera*, *Vermamoebidae*, LKM74-lineage, *Tetramitia*_VI_X, *Echinamoebidae*, *Acramoebidae*, *Rotosphaerida*_XXX, together with the protist families *Kinetoplastea*_XX and *Limnofilidae*. These taxa included both amoebae described as hosts of waterborne pathogens (Boamah et al., 2017) and bacterivorous protists (Bass et al., 2009; Kostygov et al., 2021). In prokaryotes, the highest cross-kingdom weights were observed for *Weeksellaceae* and *Micavibrionaceae*, the latter also showing a high degree of centrality. These taxa include bacteria with different ecological strategies, from resistance to amoebal digestion to bacterial predation (Kadouri et al. 2013; Lim et al. 2020). *Legionellaceae* showed a low cross-kingdom weight and an intermediate degree centrality, consistent with its associations being distributed across many ASVs (Fig. 5, Fig. S9).

### The eukaryotic taxa associated with Legionella extend beyond the known hosts

As *Legionella* bacteria showed a high prevalence throughout the sampled biofilms and are known to have close interactions with several FLAs (Boamah et al., 2017), we decided to further investigate the eukaryotes co-occurring with *Legionella* ASVs and how they influence the detection of this genus. We considered two lists of amoeba taxa: a first list included only taxa reported in the literature as hosts for *Legionella* (hereafter named as “Literature amoebae list”) and a second extending the first with taxa derived from the co-occurrence network analysis and the core community (hereafter “Integrated amoebae list”) (Table 1).

**Table 1:**
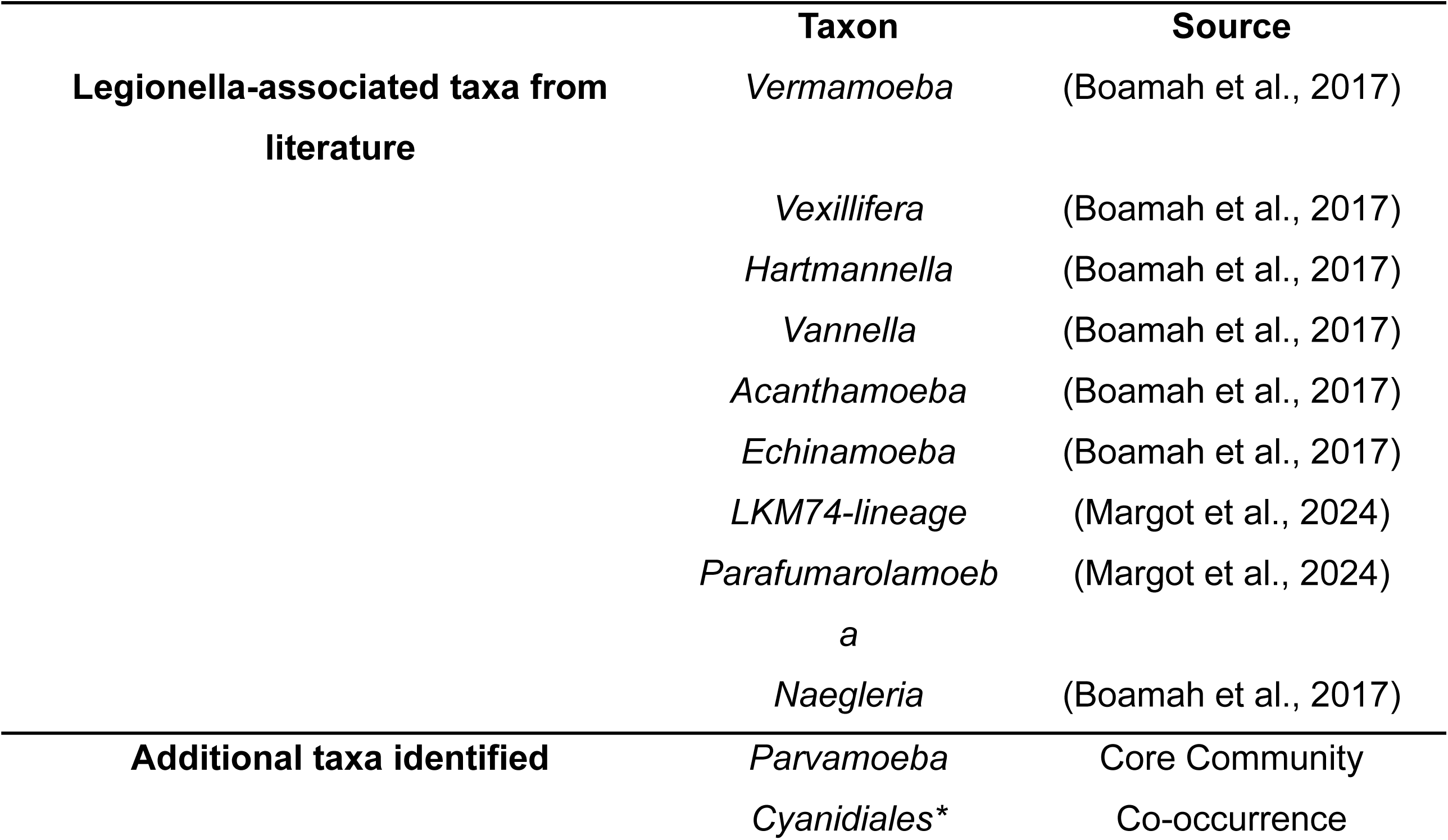
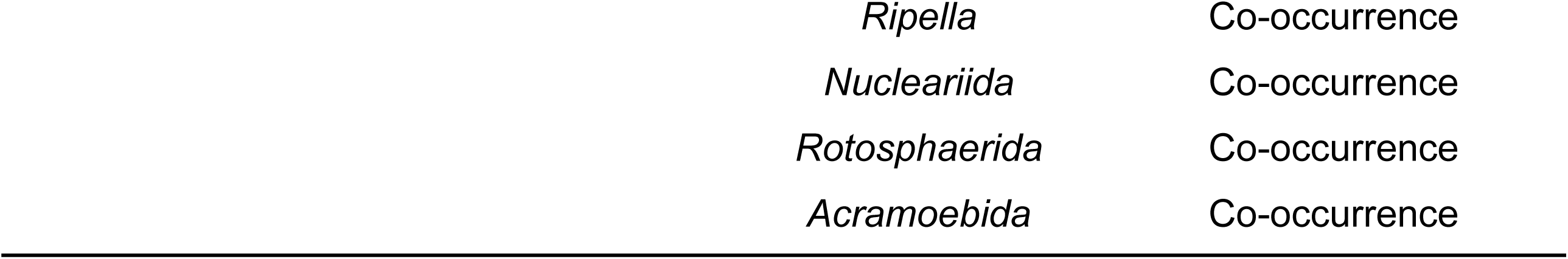
The amoebae taxa selected for further analysis either based on literature information (i.e., Literature amoebae list) or based on the additional taxa identified (i.e., Integrated amoebae list).

Comparing the richness of *Legionellaceae* ASVs to the one of the rest of the biofilm communities, we detected significant correlations with both prokaryotic (rho = 0.40, *p* = 0.003, Fig. 6a) and eukaryotic community (rho = 0.54, *p* = 0.0006, Fig. 6b). Yet, no significant correlation was found between *Legionellaceae* and the amoebae known as reservoir from literature (rho = 0.184, *p* = 0.281, Fig. S8a). On the contrary, a significant correlation emerged when considering the richness of the families within the Integrated amoebae list (rho = 0.425, *p* = 0.009, Fig. 6c), indicating that several hosts of *Legionella* exist beyond the ones recognized by scientific literature and the importance to consider them when assessing the *Legionella* environmental prevalence.

**Figure 6:**
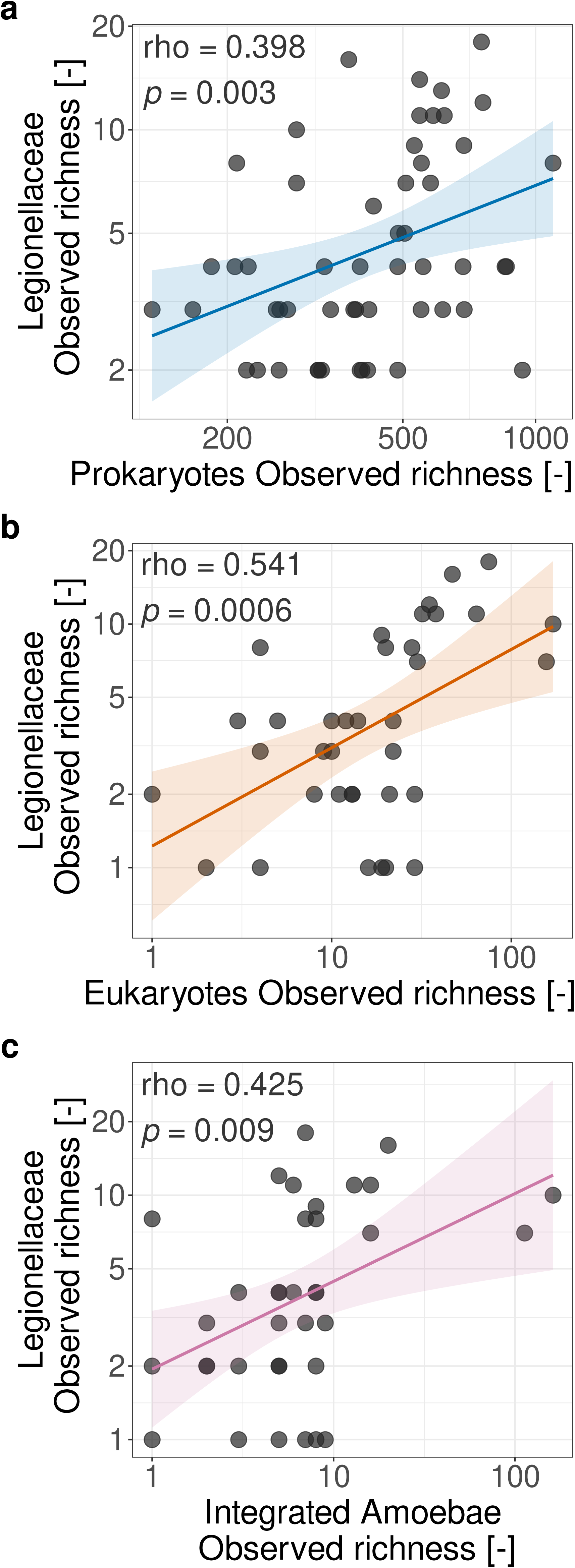
Observed richness correlations between *Legionellaceae* and microbial community diversity indices. (a) Spearman correlation between *Legionellaceae* observed richness and prokaryotes, (b) eukaryotes and (c) the integrated amoebae list, which includes both the literature-based taxa and those identified through co-occurrence network analysis (Table 1). Each point represents a sample. Axes are shown on a log10 scale. Linear regression lines with 95% confidence intervals are shown.

The cross-kingdom co-occurrence network of *Legionellaceae* ASVs did not show any network hub, suggesting that each taxa has different associations (Fig. S9). Most positive interactions between *Legionellaceae* ASVs and amoebae involved taxa within known eukaryotic families such as LKM74-lineage_X, *Parafumarolamoeba* and *Echinamoeba*, while most of the negative interactions were with unassigned taxa. Notably, none of the associations shown were between *Legionella* and *Vermamoeba*, despite being the most studied host-pathogen system in drinking water (Delafont et al., 2018). *Vermamoebidae* ASVs grouped in 4 hubs (Fig. S10) and showed associations with *Babeliaceae* and *Rickettsiales*, strictly intracellular bacteria of protists (Horn & Wagner, 2004), *Mycobacterium* and *Chryseobacterium*, both known to resist amoebal digestion (Greub & Raoult, 2004), and *Sphingomonas*. Given that these predicted interactions agree with the known ecology of these taxa, it is possible their competition for intracellular niches might have limited *Legionella* the possibility to reproduce within *Vermamoebidae,* similarly to what described for other intracellular protist bacteria (L. König et al., 2019; Maita et al., 2018).

## Discussion

In this study, we investigated the prokaryotic and eukaryotic communities of non-disinfected drinking water shower hose biofilms collected across Switzerland. We studied their composition (Fig. 1 and Fig. 2), their cross-kingdom association (Fig. 5) and the presence of potential opportunistic waterborne pathogens, such as *Legionella* (Fig. 1 and Fig. S2).

Despite originating from geographically separated locations, the shower hose biofilms shared a group of prokaryotic families detected across all samples, suggesting consistency in community membership at the family level in these environments. Several of these families have also been reported in biofilms from other non-disinfected drinking water systems, including distribution pipes (G. Liu et al., 2020) and shower hoses (Cavallaro et al., 2023; Proctor et al., 2018). *Sphingomonadaceae*, for example, has previously been identified among the dominant families in non-disinfected drinking water communities (G. Liu et al., 2014). These observations suggest similarities across non-disinfected systems, although they do not necessarily establish the presence of an overall common core community. Differences were also apparent, including the non-detection of members of the phylum *Patescibacteria* in our samples, which have frequently been reported in non-disinfected drinking water systems (Dai et al., 2020).

The disinfection regime is one of the main factors shaping drinking water microbiome composition, supporting the observations of Foucault and colleagues (Foucault et al., 2026). Comparison of our data with the global drinking water genome catalogue, which is largely derived from disinfected systems (Sudarshan et al., 2024), revealed notable differences in the representation of several genera. *Sphingomonas*, *Sphingopyxis*, *Bradyrhizobium*, OLB17, *Brevundimonas*, *Phenylobacterium*, *Caulobacter*, *Afipia*, and Ga0077523 were abundant in our shower hose biofilms but were either absent or present at only low abundance in the reference catalogue. Conversely, UBA4765, proposed as *Raskinella* (Sudarshan et al., 2024), was not detected in our shower hose biofilms. However, these differences cannot be attributed directly to disinfection practices, as the compared datasets also differ in sample type and analytical approach. In particular, the predominance of bulk water samples in the reference catalogue (Sudarshan et al., 2024) may contribute to the observed differences, given the established compositional differences between planktonic and biofilm-associated communities (G. Liu et al., 2014), as well as primer biases.

The eukaryotic core community was dominated by amoebae (Fig. 2), including taxa described as hosts for waterborne pathogens, such as *Vermamoebidae* and *Vannellidae* (Boamah et al., 2017; Crull et al., 2025; Delafont et al., 2018), and taxa proposed as potential hosts for *Legionella* spp., such as LKM74-lineage and *Vahlkampfiidae* (Margot et al., 2024). LKM74-lineage was also previously reported as highly abundant in drinking water biofilms (Cavallaro et al., 2023; Margot et al., 2024) and with a high transcriptional activity (Inkinen et al., 2019). The eukaryotic dataset had a relatively low sequencing depth and under-developed reference databases that limited taxonomic assignment. An example thereof is the clear misclassification of *Cyanidiales*_X, which phylogenetic analysis revealed to belong to the amoeboid genus of *Angulamoeba*. This limited taxonomic resolution is a recognized issue: in a similar dataset, 85% of the eukaryotic community remained unclassified at genus level, against 64% of the bacterial one (Cavallaro et al., 2023). These evidences highlight the need for a deeper eukaryotic sequencing and improved taxonomic databases to fully characterize the community in drinking water biofilms and their ecology, as pointed out also by other authors (Gabrielli et al., 2023; Inkinen et al., 2019).

Microdiversity analysis revealed that prokaryotic OTUs with greater internal phylogenetic diversity were detected across a larger proportion of samples (Fig. 1b), with total phylogenetic distance emerging as a stronger predictor of environmental prevalence than the effective number of ASVs. This suggests that phylogenetic differentiation among variants within an OTU, rather than variant richness alone, may be associated with their occurrence across different building water systems. Such a pattern is consistent with previous work linking microbial microdiversity to ecological differentiation and persistence across different buildings, such as institutional and residential buildings (He et al., 2025), and in different environmental conditions and ecosystems (García-García et al., 2019; Orta-Ponce et al., 2025). Notably, OTUs belonging to the core prokaryotic families also exhibited high within-OTU phylogenetic diversity, suggesting that microdiversity may contribute to their widespread occurrence across drinking water biofilms. These results show that shower hose biofilms from non-disinfected systems share a conserved core of prokaryotic and eukaryotic taxa and that phylogenetic diversity inside an OTU appears to contribute to the persistence of prokaryote taxa across non-disinfected drinking water biofilms.

The statistical associations between prokaryotic and eukaryotic communities (Fig. 3 and Fig. 4) showed a clear trend towards the concept of so-called *diversity-begets-diversity*. This concept was proposed by Calcagno and coworkers (Calcagno et al., 2017), demonstrated across multiple ecological niches (Hamilton et al., 2020; Madi et al., 2020) and previously observed at global scale in drinking water metagenomes (Gabrielli et al., 2023). Going in-depth of the cross-kingdom network (Fig. 5), the eukaryotic families showing the strongest associations included both well-known and poorly characterized taxa: free-living amoebae such as *Vermamoebidae* and *Echinamoebidae*, already described as hosts for waterborne pathogens (Boamah et al., 2017), *Rotosphaeridae* and *Nucleariida*, which are known to harbour endosymbiotic proteobacteria (Dyková et al., 2003), *Kinetoplastea*_XX, which includes flagellates that can be important human pathogens such as *Trypanosoma* and *Leishmania* (Kostygov et al., 2021) and *Limnofilidae*, bacterivorous cercozoan protists (Bass et al., 2009). On the prokaryotic side, taxa with different ecological strategies were detected, for example *Weeksellaceae*, including the genus *Chryseobacterium*, in which some species are able to survive amoebal digestion (Greub et al., 2004), *Legionellaceae*, which replicate within amoebal hosts (Boamah et al., 2017; Greub et al., 2004) and *Micavibrionaceae*, predatory epibiotic bacteria (Wang et al., 2011). On a fundamental level, this raises important questions on how drinking water microbes interact across trophic levels. Three relatively well-known interactions would influence cross-kingdom diversity. First, many protist species graze directly on prokaryotes as their primary food source, and several studies showed species-specific grazing habits (Bornier et al., 2021; NandaKafle et al., 2023; Weger et al., 2024), suggesting that both the act of selective grazing and defensive traits against grazing may contribute to diversity in co-existing populations. Second, a variety of symbiotic bacteria co-evolve with specific eukaryotes (Horn & Wagner, 2004; Leitsch et al., 2010; Steele et al., 2023), adding to the prokaryotic diversity in a variety of niches occupied by the host. Third, multiple bacterial species such as *Legionella* spp. requires an eukaryotic host for replication, where host specific infection (Declerck et al., 2007; Shaheen & Ashbolt, 2021) as well as defensive mechanisms of some hosts, would likely contribute to environmental diversity of both kingdoms. It is recognised that many of the possible relationships remain unresolved, and a mere statistical association is not sufficient to demonstrate a causative interaction. Notwithstanding, these results indicate that the associations between prokaryotic and eukaryotic communities reflect different types of ecological relationships, from resistance to grazing to shared bacterial prey, rather than host-pathogen interactions only. This broader context is relevant for interpreting the relationships of *Legionella*, which have so far been described mainly in terms of a small number of amoebal hosts.

*Legionella* is a bacterial genus of considerable public health relevance (ECDC, 2026), of which several sequence variants were detected in nearly all of our biofilm samples, often co-occurring with each other and in association with various eukaryotes (Fig S2, Fig. 7 and Fig. S9). Three aspects of *Legionella* diversity were particularly noticeable. First, *L. waltersii* was by far the most prevalent identified species (present in 59% of the samples). This species was first isolated from cooling tower water (Benson et al., 1996), and is only rarely associated with clinical outcomes in humans (C. König et al., 2005). Second, the species with by far the highest clinical relevance, *L. pneumophila*, was only detected in 5% of samples, suggesting that shower hoses may not necessarily be a primary niche for *Legionella* in building plumbing systems. This is seemingly corroborated by a recent source attribution study in Switzerland, which linked only a small fraction of clinical cases to samples collected from patients’ showers (Bigler et al., 2026). While some of the other species such as *L. birmighamensis, L. erythra* and *L. rubrilucens* have rarely reported clinical associations (Matsui et al., 2010; Papadakis et al., 2018; Wilkinson et al., 1987), these are generally regarded as considerably less relevant for public health than *L. pneumophila* (ECDC, 2026). Thirdly, our data showed co-occurence of different *Legionella* species in the same biofilm samples, following similar previous observations in drinking water biofilm samples (Cavallaro et al., 2023; Foucault et al., 2026; Margot et al., 2024). The majority of the *Legionella* ASVs remain, in fact, unassigned at species level, and phylogenetic analysis places most ASVs outside the clades of known cultivated species used as references. This reflects both the high intra-genus diversity of the 16S rRNA gene within *Legionella* (Hassler et al., 2022) and the incomplete coverage of reference databases for environmental *Legionella*. As an example, a recent study demonstrated that the inclusion of metagenome-assembled genomes nearly doubles the number of recognized *Legionellaceae* species with many novel species forming clades distant from currently isolated taxa (Gabrielli et al., 2024). Legionella require hosts for replication (Declerck et al., 2007; Shaheen & Ashbolt, 2021). Yet, no clear correlation was found with amoebae described in the literature as *Legionella* hosts, or specifically with the prevalent *Vermamoeba*, despite this being the one of the relationships most studied in drinking water (Delafont et al., 2018). Positive associations were found with other amoebae such as LKM74-lineage, *Parafumarolamoeba* and *Echinamoeba*. LKM74-lineage, a poorly characterized amoebozoan family, has been proposed as a potential *Legionella* host in drinking water biofilms (Margot et al., 2024), and the high prevalence in the dataset suggests that its role, in particular the relation with waterborne pathogens, deserves further investigations. Our results indicate that a broader range of eukaryotic taxa may have a role in *Legionella* persistence in non-disinfected shower hose biofilms than the ones classically described. However, considerable further research is needed to link the environmental observations from studies like our own with the potential ecological interactions to untangle observed associations. As discussed above, multiple cross-kingdom interactions can occur between *Legionella* species and protists (Hammes et al., 2025), and these interactions will depend on both the *Legionella* species present (Buse & Ashbolt, 2011) and the protist species present (Shaheen & Ashbolt, 2021). Potential outcomes can include co-existence without uptake (Shaheen & Ashbolt, 2021), uptake without replication (Bornier et al., 2021), replication where the protist host survives (Horn & Wagner, 2004) and replication where the protist hosts are destroyed (Shaheen & Ashbolt, 2021).

In conclusion, our work shows that prokaryotic and eukaryotic communities of non-disinfected shower hose biofilms are structured around a conserved core of ASVs and are associated with each other, even if the exact ecological associations remain unresolved. Eukaryotic taxa relevant for *Legionella* are likely to extend beyond those described in the literature as known hosts. Hence, characterizing the eukaryotic fraction of drinking water biofilms is therefore essential to understand the ecological context in which opportunistic pathogens persist. Ultimately, a deeper understanding of *Legionella* diversity and its relationship with protists in drinking water systems could support the development of better monitoring and management strategies in building plumbing systems.

## Methods

### Sampling and biofilm collection

128 samples and controls were obtained from participants at the SwissLegio project (Fischer, Bigler, et al., 2023), previous approval. Shower hoses were collected and kept in cooled boxes during the transport, to be refrigerated at 4 °C until processing. Biofilms were collected by cutting the shower hoses and scraping the material with the backside of a sterile 1 mL pipette tip. 2 mL of biofilm from each sample were frozen at −80 °C until processing.

### DNA extraction and quantification

The DNA was extracted using the FastDNA SPIN kit (MP Biomedicals), following an adapted protocol. 0.8 mL of scraped biofilms were incubated for 1 h at 37°C and 300 rpm with 294 µL of 1 × Tris-EDTA pH 8.0 (100 mM Tris Base, 10 mM EDTA, G-Bioscience) and 6 µL of lysozyme (50 mg/mL, Thermo Scientific). After incubation, 300 µL of CLS-TC buffer (FastDNA SPIN kit, MP Biomedicals) and 30 µL of proteinase K (20 mg/mL, Thermo Fisher Scientific) were added to the samples. Samples were then incubated for 30 min at 56°C, 300 rpm. Lysing matrix and 630 µL chloroform/Isoamyl alcohol (ratio 24:1, Brunschwig) were added to the incubated samples. After 5 min of vortexing, samples were centrifuged for 10 min at 4 °C and 14,000 × g and 750 µL of the Binding Matrix (FastDNA SPIN kit, MP Biomedicals) added to the upper aqueous phase. The next steps followed the manufacturer’s protocol.

Negative and positive controls were processed during each extraction batch. Negative controls were clean filters, while positive were filters previously used for *L. pneumophila* culture filtration. Mock communities for both prokaryotes and eukaryotes were added using ZymoBIOMICS Microbial Community DNA Standard II Log Distribution (Zymo Research), ABRF-MGRG 10 Strain Even Mix Genomic Material (ATCC) and Mycobiome Genomic DNA Mix (ATCC).

### Amplicon sequencing and processing

Prokaryotic and eukaryotic libraries were sequenced at the Next Generation Sequencing (NGS) Platform of the University of Bern on a PacBio Revio instrument. Reads processing was performed on the Euler high-performance computing cluster at ETH Zürich, Switzerland.

### Prokaryotic community profiling

Library preparation for full-length HiFi 16S amplicon sequencing was performed using the Kinnex library preparation method (Al’Khafaji et al., 2024) and the 27F (AGAGTTTGATCMTGGCTCAG) and 1492R (TACCTTGTTACGACTT) primers (Frank et al., 2008). Raw reads were processed using R v.4.5.3 and RStudio v.2024.01.2+418 (Posit team, 2024; R Core Team, 2026). Primers were removed and reads filtered using the DADA2 v.1.40.0 (Callahan et al., 2016) functions *removePrimers* and *filterAndTrim* (minLen = 1200, maxLen = 1600, maxEE = 2). *learn_errors* function was adapted to PacBio Revio reads with binned Phred scores (Callahan, n.d.; Callahan et al., 2019). The resulting error model was used to denoise the ASVs with *dada* function. Chimeras were removed with consensus method and minFPOA = 8. Taxonomy was assigned using the Naive Bayes classifier algorithm implemented in DADA2 (Callahan et al., 2016), using the *assignTaxonomy* function and the GTDB R220 database (Parks et al., 2026). ASVs were clustered at species-level using a threshold 98.7% of similarity (Yarza et al., 2014) using the *Clusterize* function from DECIPHER v.3.8.0 (Wright, 2024). Finally, the prokaryotic dataset was, then, rarefied using the function *rarefy_even_depth* from phyloseq v.1.56.0 (McMurdie & Holmes, 2013) at 35000 reads.

### Eukaryotic community profiling

Long-read amplicon HiFi sequencing of the 18S-ITS-28S region was performed using the TAReuk454FWD1 (CCAGCASCYGCGGTAATTCC) and 21R (GACGAGGCATTTGGCTACCTT) primers (Latz et al., 2022). Raw reads were processed using the NextITS pipeline v.0.5.0 (Mikryukov et al., 2026), omitting the ITS extraction step (--its_region none). Denoising and clustering were performed using the UNOISE3 algorithm (--unoise true). The resulting ASVs were further curated by identifying 18S rRNA sequences using barrnap v.1.10.5 (Seemann, n.d.) and discarding sequences with none or more than one predicted rRNA. ASVs taxonomy was assigned based on the predicted 18S rRNA sequences using the Naive Bayes classifier algorithm implemented in DADA2 (Callahan et al., 2016) relying on the PR2 database v.5.1.1 (Guillou et al., 2013). Sequences not corresponding to Eukarya have been removed. A final decontamination was performed using the decontam v.1.32.0 (prevalence method) based on the sequenced negative controls (Davis et al., 2018). The *Clusterize* function from DECIPHER was used to group the ASVs in OTUs with 98.7% of similarity. The eukaryotic dataset was, then, rarefied using the function *rarefy_even_depth* at 3000 reads.

### Phylogenetic analyses

Maximum likelihood phylogenetic trees were constructed for both datasets. For the prokaryotic rRNA dataset, ASV sequences were aligned using MAFFT v.7.525 with automatic algorithm selection (--auto) (Katoh et al., 2002) and trimmed using trimAl (-automated1) (Capella-Gutiérrez et al., 2009). The phylogenetic tree was built using IQ-TREE2 v2.2.2.6 with ModelFinder Plus (-m MFP, -mset GTR), 1000 ultrafast bootstrap replicates (-B 1000) and branch length optimization (-bnni) (Minh et al., 2020). For the eukaryotic rRNA dataset, the same alignment and trimming procedure was applied and the phylogenetic tree was constructed using FastTree v2.1.11 with -nt -gtr parameters (Price et al., 2009). ITOL v.7.5.1 (Letunic & Bork, 2024) was used to visualize both trees. Further phylogenetic analyses were performed for specific clades of interest. Eukaryotic ASVs classified as *Cyanidiales*_XX in the eukaryotic dataset were selected to understand the presence of this taxa. Sequences were extracted and compared using BLASTn (Zhang et al., 2000) against NCBI (accessed on: March 2026). Since the BLAST results highlighted a high similarity of the *Cyanidiales*_XX ASVs with *Angulamoeba* sequences, taxa belonging to the family *Cyanidiales* (i.e., genera *Cyanidium*, *Cyanidioschyzon*, *Cyanidiococcus*) and from the genus *Angulamoeba* were selected for further phylogenetic analyses. *Galdieria maxima*, the previous classification of *Cyanidiococcus yangmingshanensis*, was also included in the selected taxa, while *Saccharomyces cerevisiae* was included as an outgroup. 18S rRNA sequences of the selected taxa were either downloaded from NCBI or, where no sequences were available, barrnap was used to predict the 18S rRNA on the whole genome sequence and the best predicted sequence was retained. Further phylogenetic analyses were also conducted on ASVs belonging to the genus *Legionella* to verify species-level taxonomic assignment obtained through GTDB R220 database. Full-length 16S rRNA sequences of known *Legionella* species (n=65) were retrieved from NCBI and used as reference sequences for phylogenetic comparison with the dataset ASVs. In addition, the 16S rRNA sequence of *Coxiella burnetii* was selected as an outgroup, being the closest taxa outside *Legionellaceae* family (Legionellales order). Sequence alignment and tree construction for *Cyanidiales*_XX and *Legionellaceae* ASVs were carried out as performed for the eukaryotic ASVs.

### Core communities assessment

ASVs belonging to the core eukaryotic and prokaryotic communities were calculated as proposed previously (Shade & Stopnisek, 2019). Briefly, ASVs-level taxa were ranked by a weighted occupancy-abundance index, and taxa were added iteratively to calculate the mean of Bray-Curtis similarity across samples. To identify the membership in the core community, a cut-off was defined using the last 2% increase method, identifying the point where additional taxa contribute marginally to the community dissimilarity. Below the point, ASVs were classified as belonging to the satellite community.

### Diversity analyses

Alpha diversity was estimated using observed richness and Shannon diversity indices, using the *estimate_richness* function from phyloseq. Beta diversity was estimated using Bray-Curtis and Jaccard dissimilarities calculated using the *distance* function from phyloseq. The presence of effects due to distinct water sources on community composition was tested using Permutational Multivariate Analysis of Variance analysis (PERMANOVA) with 9999 permutations, as implemented in the vegan R package. Cross-kingdom relationships between prokaryotes and eukaryotes were assessed using Procrustes analysis, Mantel test and PERMANOVA, based on both Bray-Curtis and Jaccard dissimilarities. Procrustes analyses and Mantel tests were performed using the *procrustes, protest* and *mantel* (Pearson correlation) functions from vegan with 9999 permutations. PERMANOVA (9999 permutations) was used to measure the fraction of variance of prokaryotic and eukaryotic communities explained by either the composition of the other community and abiotic parameters (i.e., cold water, hot water and room temperatures). Vectors describing the community composition to use in the PERMANOVA analysis were obtained by projecting community compositions on a one-dimensional dimension through Principal Coordinates Analysis (PCoA) and Non-metric Multidimensional Scaling (NMDS), and selecting the projection providing a higher Spearman correlation with the original dissimilarity matrix, similarly to what described by Badra et colleagues (Badra et al., 2025).

### Network analysis

Network analyses combining both prokaryotic and eukaryotic datasets were performed using SPIEC-EASI v.2.0.0 (Kurtz et al., 2015) using the Meinshausen-Bühlmann (MB) neighbourhood selection method and the following parameters: lambda.min.ratio = 5e-2, nlambda = 40, selection criterion = bstars, 100 repetitions and random seed = 10010. Network visualization was performed using the visNetwork R package. ASVs were filtered based on total reads count in the ASV table. Different thresholds were applied to each dataset, 5 reads for the prokaryotic dataset and more than 3 reads for the eukaryotic, to balance the lower sequencing depth of the eukaryotes dataset.

### Microdiversity and OTU-level phylogenetic diversity

Microdiversity was performed on both datasets at OTU level (98.7% similarity), similarly to what described before (García-García et al., 2019). Each OTU was rarefied at 5000 reads using the *rrarefy* function from vegan R package retaining 631 prokaryotic OTUs and 34 eukaryotic ones. Microdiversity was then calculated as the exponential of the Shannon diversity of the ASVs within each OTU, obtaining the effective number of ASVs, while the total cophenetic distance was computed as the sum of pairwise phylogenetic distances between all the ASVs. OTU prevalence was defined as the proportion of samples in which the OTU was detected after rarefaction. Microdiversity and total cophenetic distance were tested as predictors of OTU persistence using a Random Forest regression model (Breiman, 2001) with randomForest v.4.7.1.2 (ntree = 1000). Wilcoxon rank-sum test was used to assess differences of the prevalence of OTUs with microdiversity above and below the threshold of 2 effective ASVs.

## Data availability

Raw sequencing reads will be available upon publication on the European Nucleotide Archive (ENA) under the BioProject PRJEB128225.

## Acknowledgments

The Authors want to acknowledge the support of Antonia Eichelberg and Natasha Roser for their help in biofilm collection. In addition, the Authors acknowledge Eawag’s support through discretionary funding and Copan Italia S.p.A., which funded the PhD grant of Laura Caligaris, in accordance with DM 117 2/03/2023 PNRR, Missione 4.

## Declaration of AI use

Claude was used by LC to assist with coding solutions. ChatGPT (OpenAI) was used by FH to assist with language editing, clarity, paragraph structure, and the logical organization of scientific arguments in selected excerpts from the first section of the Discussion. Neither the full manuscript nor underlying research datasets were uploaded. No citations were generated. All suggested code solutions and revisions were reviewed by the authors, who retained full responsibility for the manuscript’s content and scientific interpretations.

