## Supplementary information for "Integrating 16S and 18S sequencing reveals coupled prokaryotic and eukaryotic relationships in drinking water biofilms"

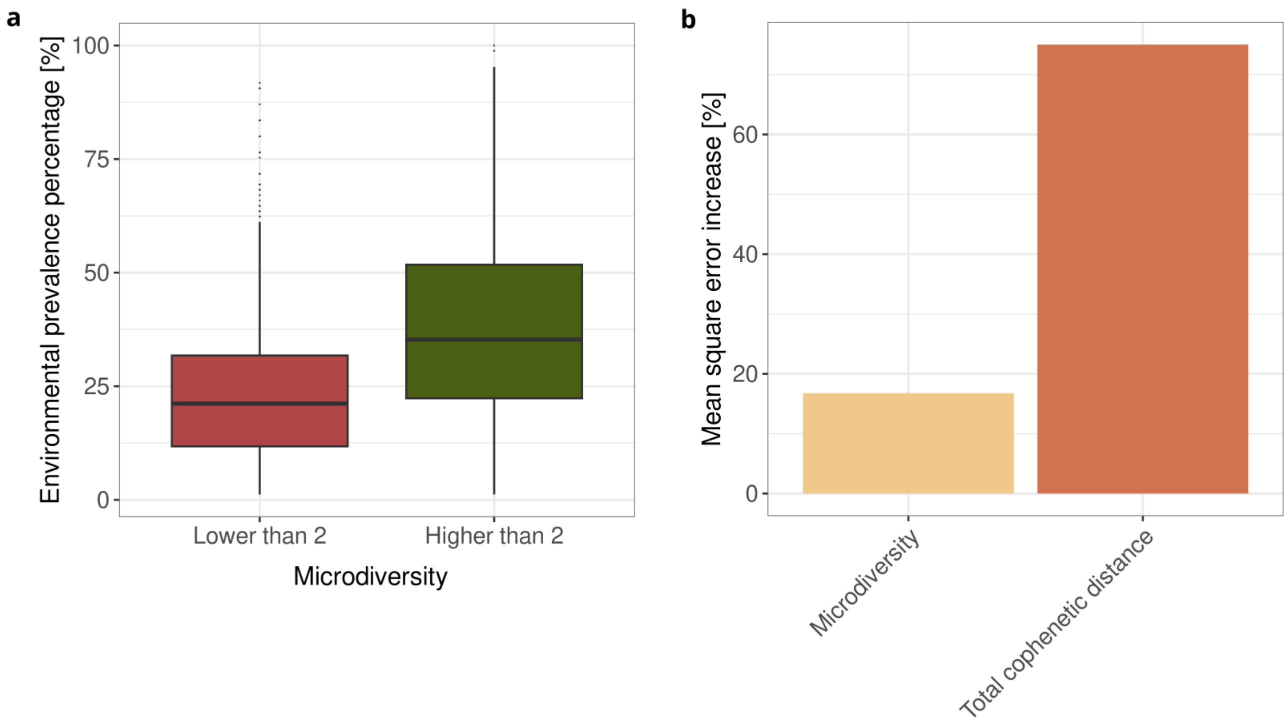


**Supplementary Figure 1.** Microdiversity and detention frequency for the prokaryotic dataset. Boxplots show the detention percentage for OTUs with microdiversity lower or higher than 2 effective ASVs (a), while barplots show the random forest variable importance based on mean square error increase when microdiversity or total cophenetic distance is removed from the model (total cophenetic distance 76% against 15.4% for microdiversity)(b).

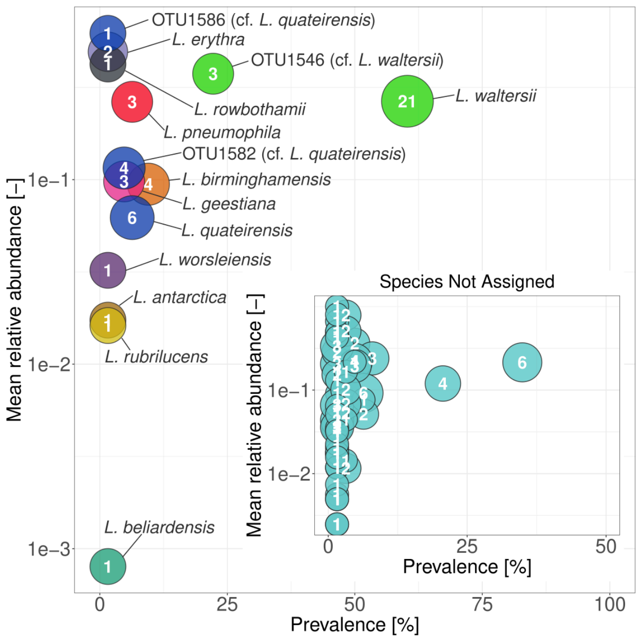


**Supplementary Figure 2: Prevalence and mean relative abundance of *Legionella* species identified across samples.** Each circle represents a species-level or taxa with 98.7% of identity; the number inside each circle indicates the number of ASVs assigned to that taxon. Circle size is proportional to the number of ASVs. Taxa labeled as "cf." indicate OTU98.7 clusters phylogenetically related to the indicated species but without formal species assignment.

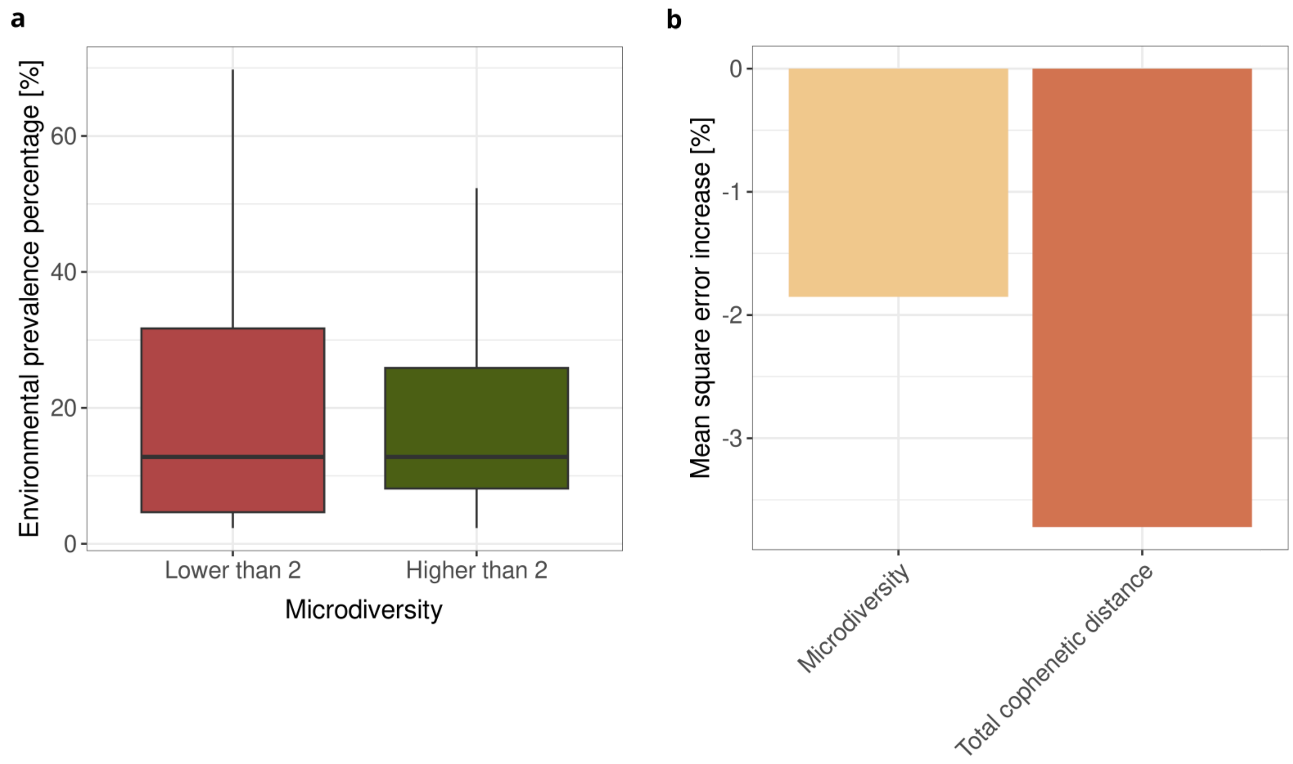


**Supplementary Figure 3.** Microdiversity and detention frequency for the eukaryotic dataset. Boxplots show the detention percentage for OTUs with microdiversity lower or higher than 2 effective ASVs (a), while barplots show the random forest variable importance based on mean square error increase when microdiversity or total cophenetic distance is removed from the model (total cophenetic distance -2.784% against 0.098% for microdiversity)(b).

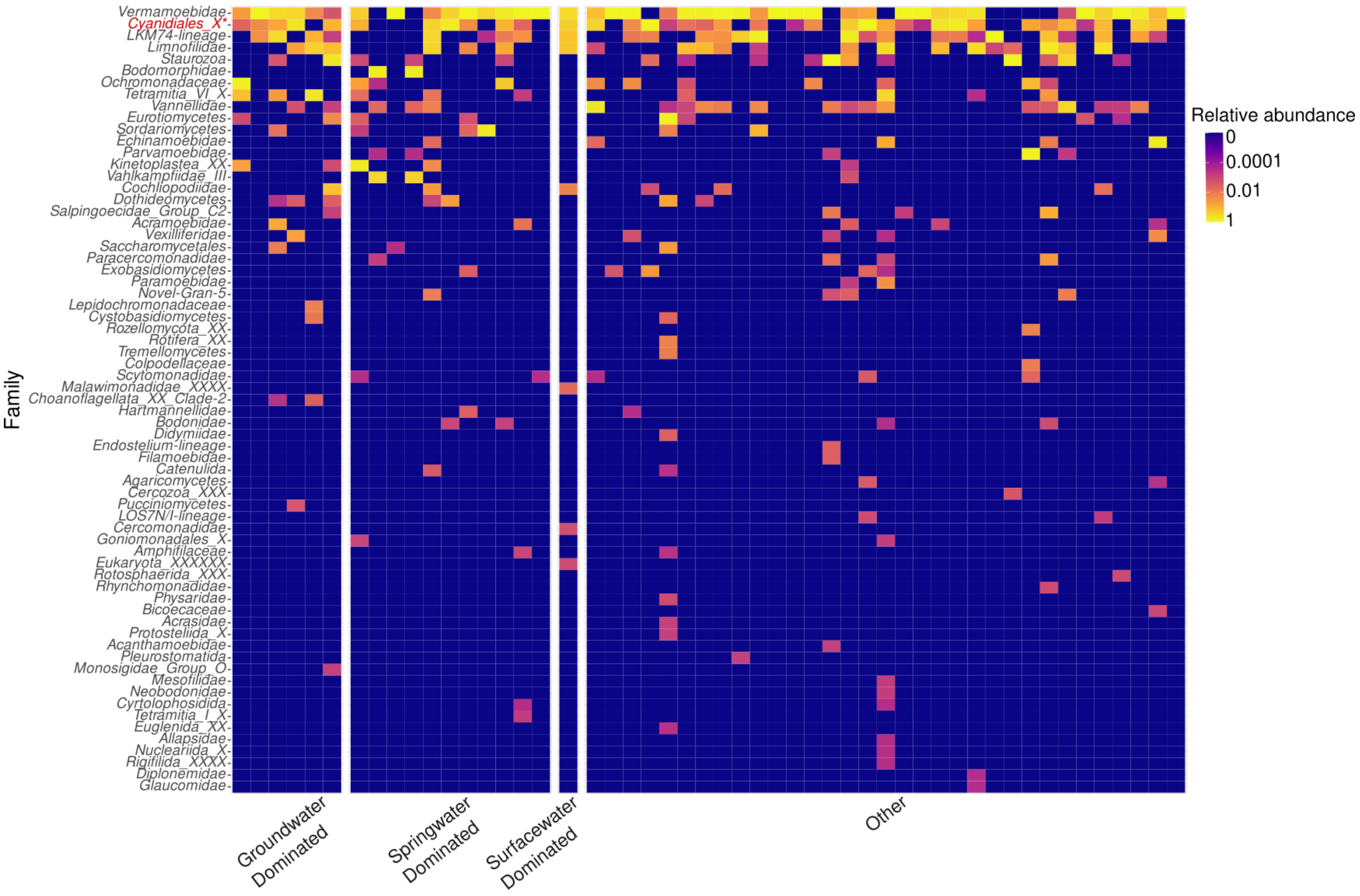


**Supplementary Figure 4:** Relative abundance of eukaryotic families across samples grouped by water source. Each row represents a family-level taxon and each column a sample. Color scale indicates mean relative abundance on a log10 scale. Samples are grouped by water source:

Groundwater Dominated, Springwater Dominated, Surfacewater Dominated and Other. Asterisk and red color indicate a putative misclassification (see main text).

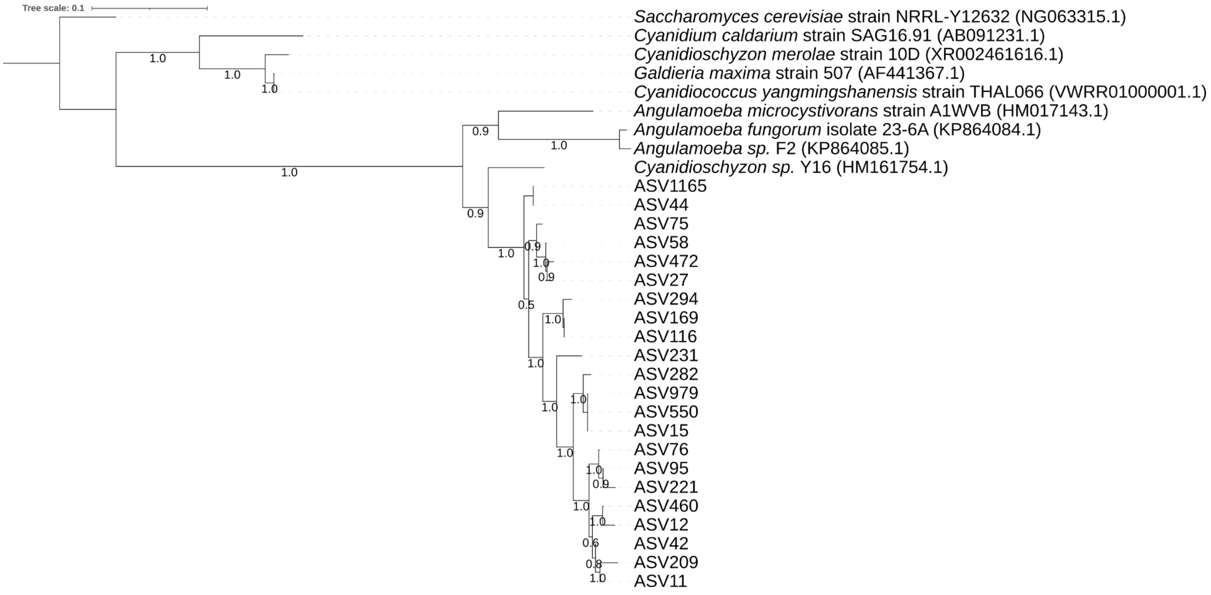


**Supplementary Figure 5:** Maximum likelihood phylogenetic tree of ASVs assigned to Cyanidiales_X using the PR2 v5.1.1 database. Reference sequences from the most closely related families identified by BLAST are included for comparison. *Saccharomyces cerevisiae* strain NRRL-Y12632 is used as outgroup; *Galdieria maxima* strain 507 is the old classification for *Cyanidiococcus yangmingshanensis*.

| ASV name | Top Hit | Accession Number | Identity (%) | First classified hit | Accession Number | Identity (%) |
|---|---|---|---|---|---|---|
| ASV11 | *Angulamoeba* sp. FN806 | KP864086.1 | 97.29 | - | - | - |
| ASV12 | *Angulamoeba* sp. FN806 | KP864086.1 | 95.63 | - | - | - |
| ASV15 | *Angulamoeba* sp. FN806 | KP864086.1 | 100 | - | - | - |
| ASV27 | Uncultured eukaryote | AB695530.1 | 96.52 | *Angulamoeba* sp. FN806 | KP864086.1 | 95.5 |
| ASV42 | *Angulamoeba* sp. FN806 | KP864086.1 | 96.75 | - | - | - |
| ASV44 | Uncultured eukaryote | AB695530.1 | 97.22 | *Angulamoeba* sp. FN806 | KP864086.1 | 95.96 |
| ASV58 | Uncultured eukaryote | AB695530.1 | 97.06 | *Angulamoeba* sp. FN806 | KP864086.1 | 96.11 |
| ASV75 | Uncultured eukaryote | AB695530.1 | 97.3 | *Angulamoeba* sp. FN806 | KP864086.1 | 97.3 |
| ASV76 | *Angulamoeba* sp. FN806 | KP864086.1 | 96.23 | - | - | - |
| ASV95 | *Angulamoeba* sp. FN806 | KP864086.1 | 96.23 | - | - | - |
| ASV116 | Uncultured eukaryote | AB695530.1 | 95.79 | *Angulamoeba* sp. FN806 | KP864086.1 | 95.37 |
| ASV169 | Uncultured eukaryote | AB695530.1 | 95.79 | *Angulamoeba* sp. FN806 | KP864086.1 | 95.37 |
| ASV209 | *Angulamoeba* sp. FN806 | KP864086.1 | 95.84 | - | - | - |

| ASV name | Top Hit | Accession Number | Identity (%) | First classified hit | Accession Number | Identity (%) |
|---|---|---|---|---|---|---|
| ASV11 | *Angulamoeba* sp. FN806 | KP864086.1 | 97.29 | - | - | - |
| ASV12 | *Angulamoeba* sp. FN806 | KP864086.1 | 95.63 | - | - | - |
| ASV15 | *Angulamoeba* sp. FN806 | KP864086.1 | 100 | - | - | - |
| ASV27 | Uncultured eukaryote | AB695530.1 | 96.52 | *Angulamoeba* sp. FN806 | KP864086.1 | 95.5 |
| ASV221 | *Angulamoeba* sp. FN806 | KP864086.1 | 96 | - | - | - |
| ASV231 | *Angulamoeba* sp. FN806 | KP864086.1 | 95.32 | - | - | - |
| ASV282 | *Angulamoeba* sp. FN806 | KP864086.1 | 99.06 | - | - | - |
| ASV294 | Uncultured eukaryote | AB695530.1 | 96.16 | *Angulamoeba* sp. FN806 | KP864086.1 | 94.9 |
| ASV460 | *Angulamoeba* sp. FN806 | KP864086.1 | 96.3 | - | - | - |
| ASV472 | Uncultured eukaryote | AB695530.1 | 97.07 | *Angulamoeba* sp. FN806 | KP864086.1 | 95.97 |
| ASV550 | *Angulamoeba* sp. FN806 | KP864086.1 | 100 | - | - | - |
| ASV979 | *Angulamoeba* sp. FN806 | KP864086.1 | 100 | - | - | - |
| ASV1165 | Uncultured eukaryote | AB695530.1 | 97.22 | *Angulamoeba* sp. FN806 | KP864086.1 | 95.96 |

**Supplementary Table 1:** BLASTn results for ASVs assigned to *Cyanidiales_X* by the PR2 database. Query coverage was 98% for all ASVs. Where the top hit was an uncultured or unclassified eukaryote sequence, the first taxonomically assigned hit is also reported. Dashes indicate that the top hit was already a taxonomically identified sequence.

| Variable 1 | Variable 2 | Method | Distance | r | *p* |
|---|---|---|---|---|---|
| Prokaryotes | Eukaryotes | Procrustes | Bray-Curtis | 0.318 | 0.028* |
| Prokaryotes | Eukaryotes | Procrustes | Jaccard | 0.219 | 0.268 |
| Prokaryotes | Eukaryotes | Mantel | Bray-Curtis | 0.088 | 0.045* |
| Prokaryotes | Eukaryotes | Mantel | Jaccard | 0.057 | 0.163 |
| Legionellaceae | Eukaryotes | Procrustes | Bray-Curtis | 0.210 | 0.123 |
| Legionellaceae | Eukaryotes | Procrustes | Jaccard | 0.197 | 0.176 |
| Legionellaceae | Eukaryotes | Mantel | Bray-Curtis | 0.058 | 0.032* |
| Legionellaceae | Eukaryotes | Mantel | Jaccard | 0.049 | 0.106 |
| Legionellaceae | Eukaryotes | Procrustes | Bray-Curtis | 0.186 | 0.494 |
| Legionellaceae | Eukaryotes | Procrustes | Jaccard | 0.136 | 0.787 |
| Legionellaceae | Eukaryotes | Mantel | Bray-Curtis | 0.064 | 0.107 |
| Legionellaceae | Eukaryotes | Mantel | Jaccard | 0.0004 | 0.464 |
| Legionellaceae | Amoebae list | Procrustes | Bray-Curtis | 0.190 | 0.487 |
| Legionellaceae | Amoebae list | Procrustes | Jaccard | 0.262 | 0.168 |
| Legionellaceae | Amoebae list | Mantel | Bray-Curtis | 0.045 | 0.192 |
| Legionellaceae | Amoeba list | Mantel | Jaccard | -0.085 | 0.951 |
| Legionellaceae | Amoebae integrated list | Procrustes | Bray-Curtis | 0.9724 | 0.605 |
| Legionellaceae | Amoebae integrated list | Procrustes | Jaccard | 0.984 | 0.836 |
| Legionellaceae | Amoebae integrated list | Mantel | Bray-Curtis | 0.060 | 0.123 |
| Legionellaceae | Amoebae integrated list | Mantel | Jaccard | -0.075 | 0.913 |
| Amoebae list | Prokaryotes | Procrustes | Bray-Curtis | 0.9004 | 0.031* |
| Amoebae list | Prokaryotes | Procrustes | Jaccard | 0.950 | 0.254 |
| Amoebae list | Prokaryotes | Mantel | Bray-Curtis | 0.085 | 0.054 . |
| Amoebae list | Prokaryotes | Mantel | Jaccard | 0.054 | 0.192 |

| | | | | | |
|---|---|---|---|---|---|
| Amoebae integrated list | Prokaryotes | Procrustes | Bray-Curtis | 0.872 | 0.008* |
| Amoebae integrated list | Prokaryotes | Procrustes | Jaccard | 0.953 | 0.294 |
| Amoebae integrated list | Prokaryotes | Mantel | Bray-Curtis | 0.085 | 0.053 . |
| Amoebae integrated list | Prokaryotes | Mantel | Jaccard | 0.013 | 0.402 |

**Supplementary Table 2:** Mantel test and procrustes analysis on beta diversity matrices using Bray-Curtis distance and Jaccard distance (* $p < 0.05$, . $p < 0.1$).

| Variable 1 | Variable 2 | $R^2$ | F | *p* |
|---|---|---|---|---|
| Eukaryotes | Prokaryotes (PCoA axis1) | 0.075 | 2.28 | 0.026* |
| Prokaryotes | Eukaryotes (PCoA axis1) | 0.046 | 1.34 | 0.016* |
| Eukaryotes | hot water temperature | 0.056 | 1.65 | 0.090 . |
| Eukaryotes | cold water temperature | 0.02 | 0.58 | 0.874 |
| Eukaryotes | bathroom temperature | 0.025 | 0.73 | 0.708 |
| Prokaryotes | hot water temperature | 0.032 | 0.94 | 0.682 |
| Prokaryotes | cold water temperature | 0.033 | 0.96 | 0.596 |
| Prokaryotes | bathroom temperature | 0.039 | 1.13 | 0.142 |

**Supplementary Table 3:** Permutational Multivariate Analysis of Variance using Matrices across prokaryotes and eukaryotes. The distance used to calculate the matrices was Bray-Curtis (* $p < 0.05$, . $p < 0.1$).

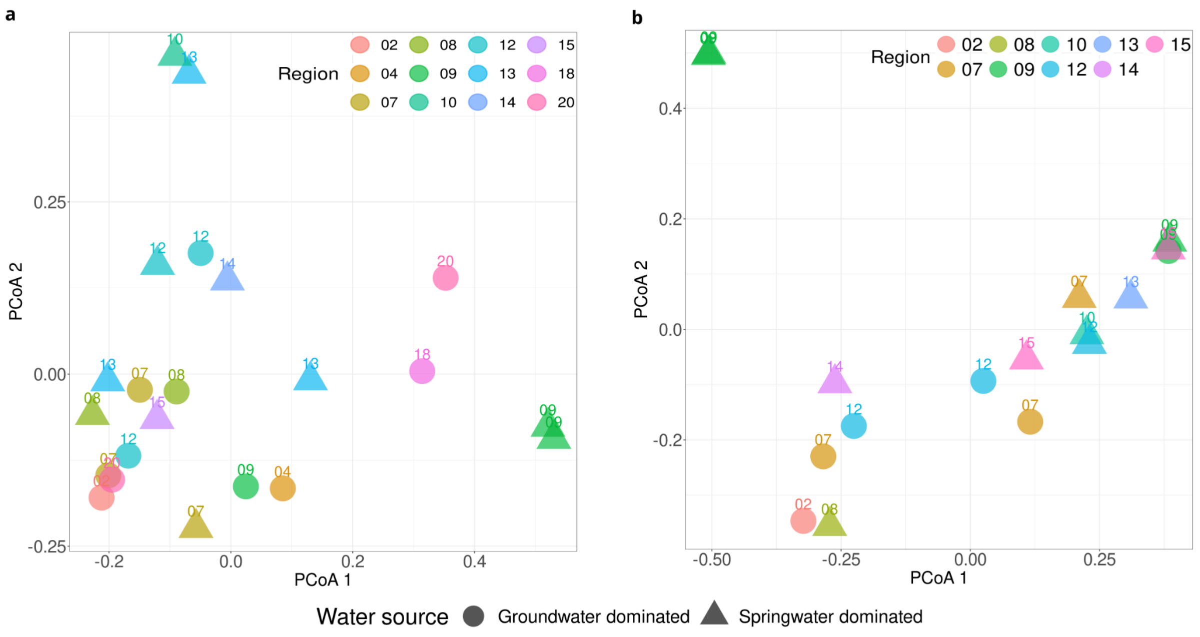


**Supplementary Figure 6:** Beta diversity of groundwater- and springwater-dominated samples based on Prokaryote (a) and Eukaryote (b) datasets. Principal Coordinates Analysis (PCoA) of Bray-Curtis dissimilarity matrices are shown for both groundwater and springwater samples. Each point or triangle represents a sample from ground or springwater; colors and labels indicate the region of origin. Samples from the same region share the same color and label number.

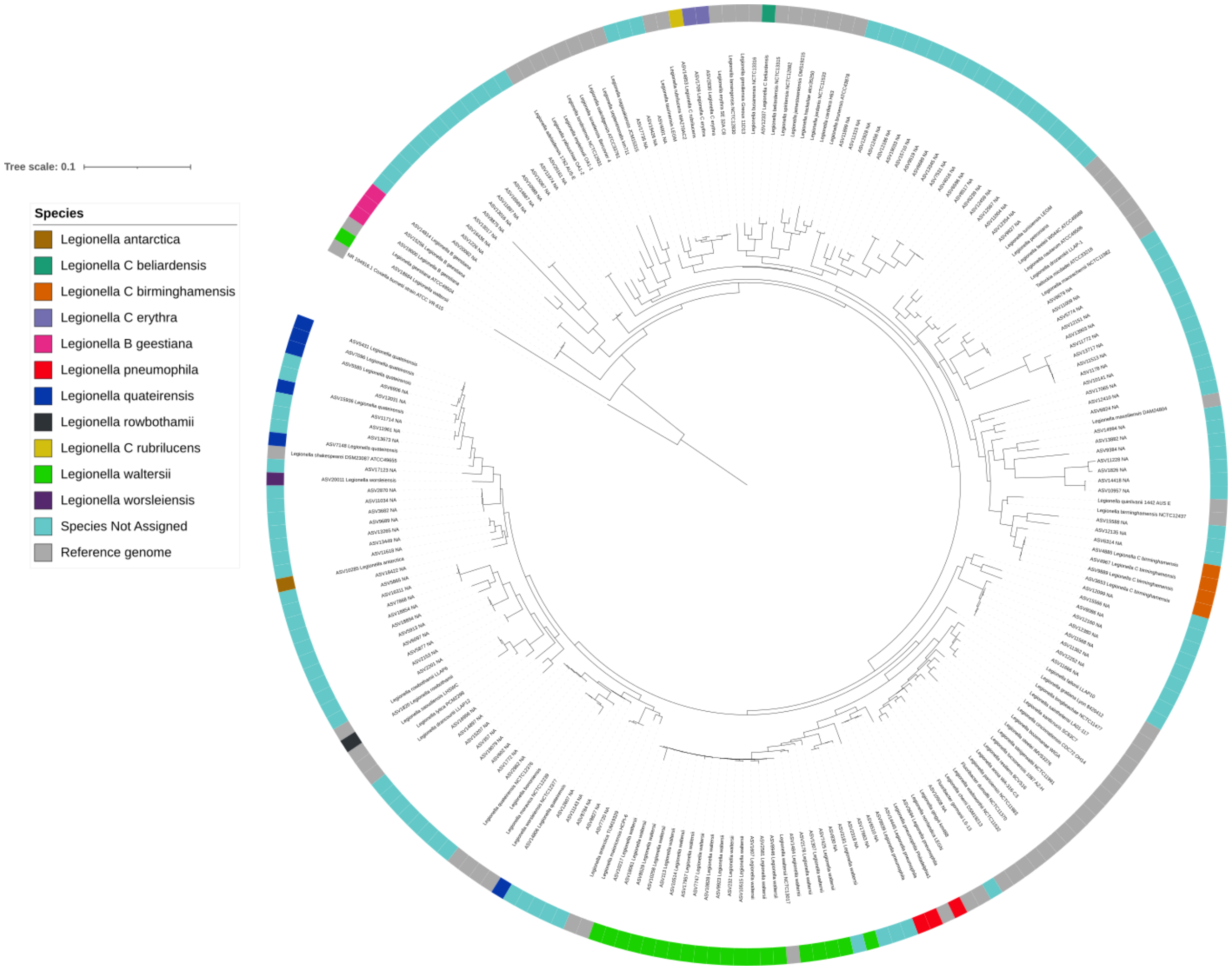


**Supplementary Figure 7:** Maximum likelihood phylogenetic tree of *Legionella* ASVs, 16S rRNA gene reference strains of the known *Legionella* species (n=65) and 16S rRNA gene of *Coxiella burnetii* (Legionellales order), used as outgroup to re-root the tree. ASVs without species assignation are shown in light blue, while the assigned ASVs share the color with the correspondent strain; in grey the outgroup and the reference strains absent in our dataset.

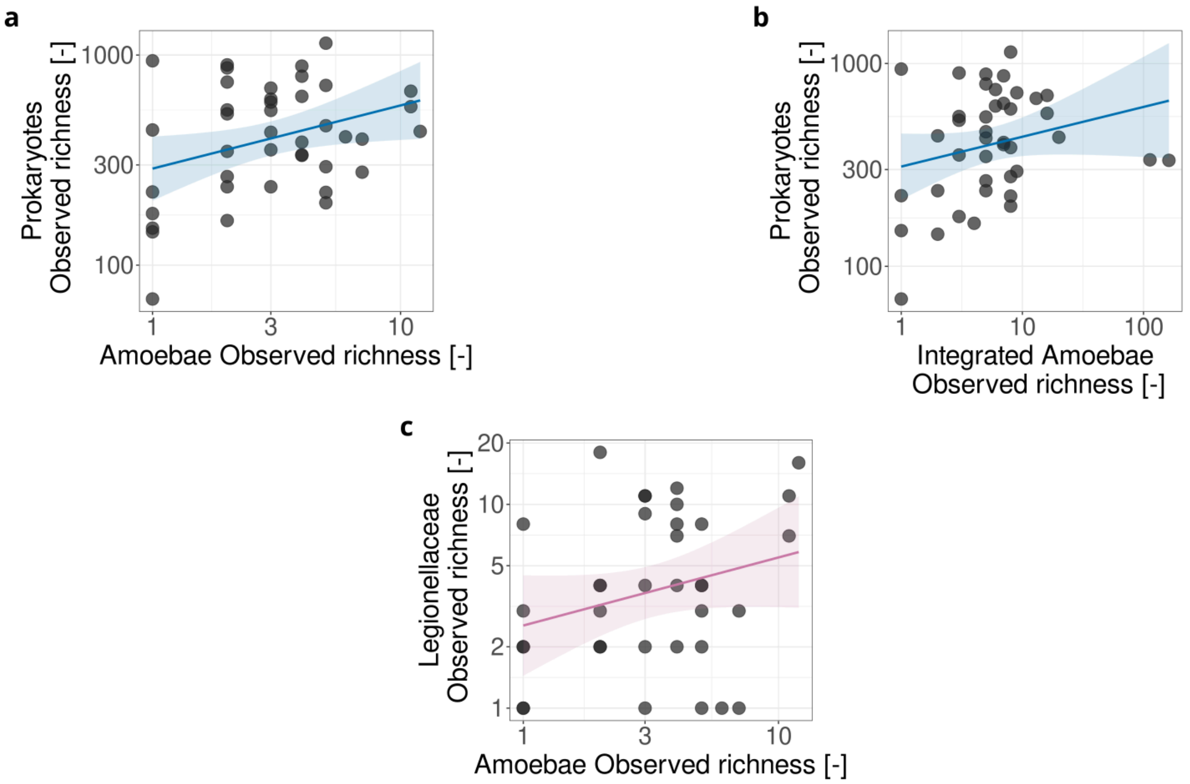


**Supplementary Figure 8:** Observed richness Spearman correlations involving the amoebae list reported in Table1. Correlations between the prokaryotic dataset and the amoebae known as reservoirs from literature (a; rho = 0.349, *p* = 0.025), between the prokaryotic dataset and the integrated amoebae list, which includes both the literature-based taxa and those identified through co-occurrence network analysis (b; rho = 0.180, *p* = 0.258) and between Legionellaceae and the amoebae known as reservoirs from literature (c) (rho = 0.184, *p* = 0.281). Each point represents a sample. Axes are shown on a log10 scale. Regression lines with 95% confidence intervals are shown.

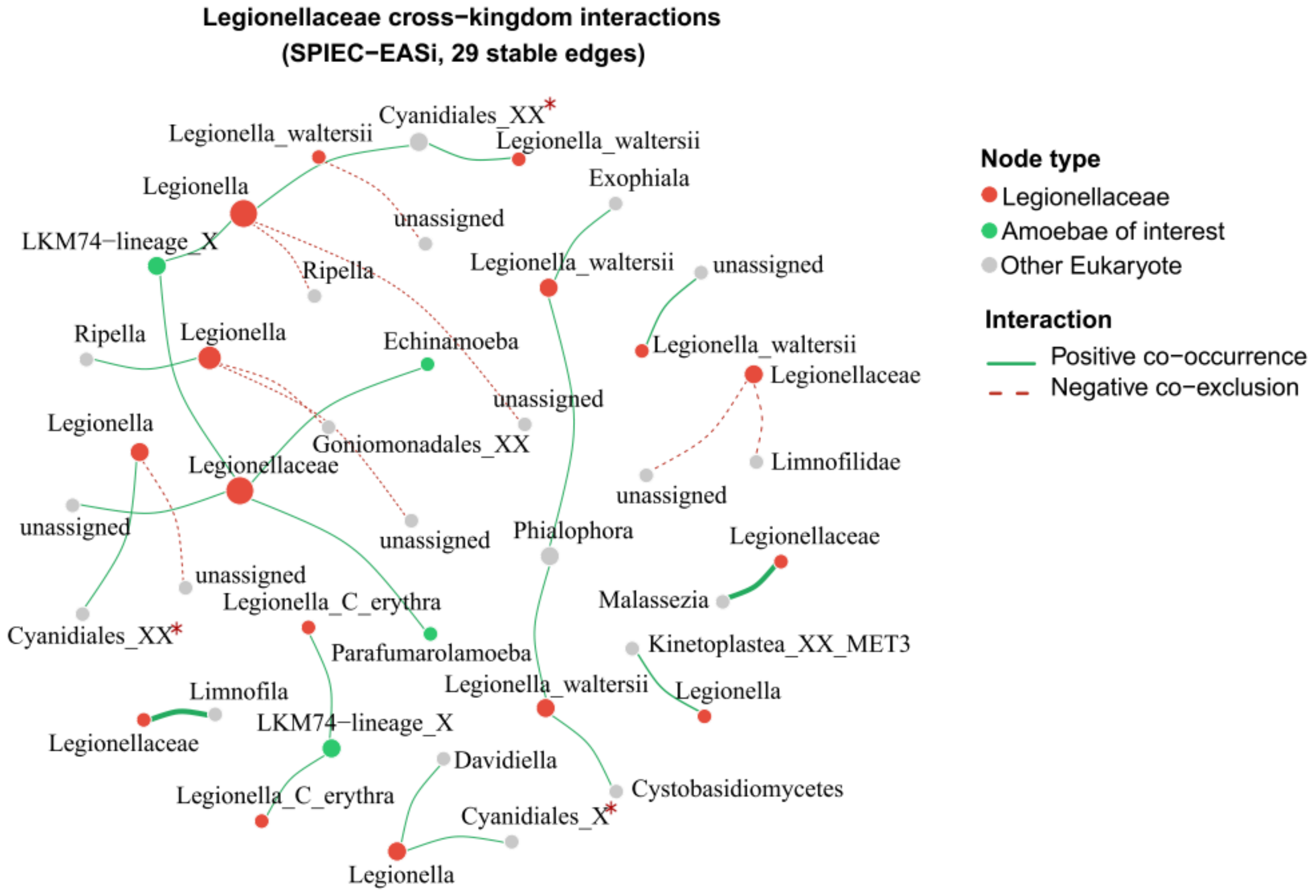


**Supplementary Figure 9:** Cross-kingdom co-occurrence network of Legionellaceae ASVs and their eukaryotic partners, inferred with SPIEC-EASI. Only stable edges identified by the StARS stability criterion are shown (n = 29). Node size is proportional to the number of stable edges. Legionellaceae ASVs are colored in red and labeled at species level where taxonomic assignment was available, or at genus/family level otherwise. Eukaryotic nodes are colored according to taxonomic group: amoebae with known associations as potential Legionella reservoirs in green, and other eukaryotes in grey. Eukaryotes labels are at family level. Red asterisk is for Cyanidiales misclassification. Green edges indicate positive co-occurrence; red dashed edges indicate negative co-exclusion.

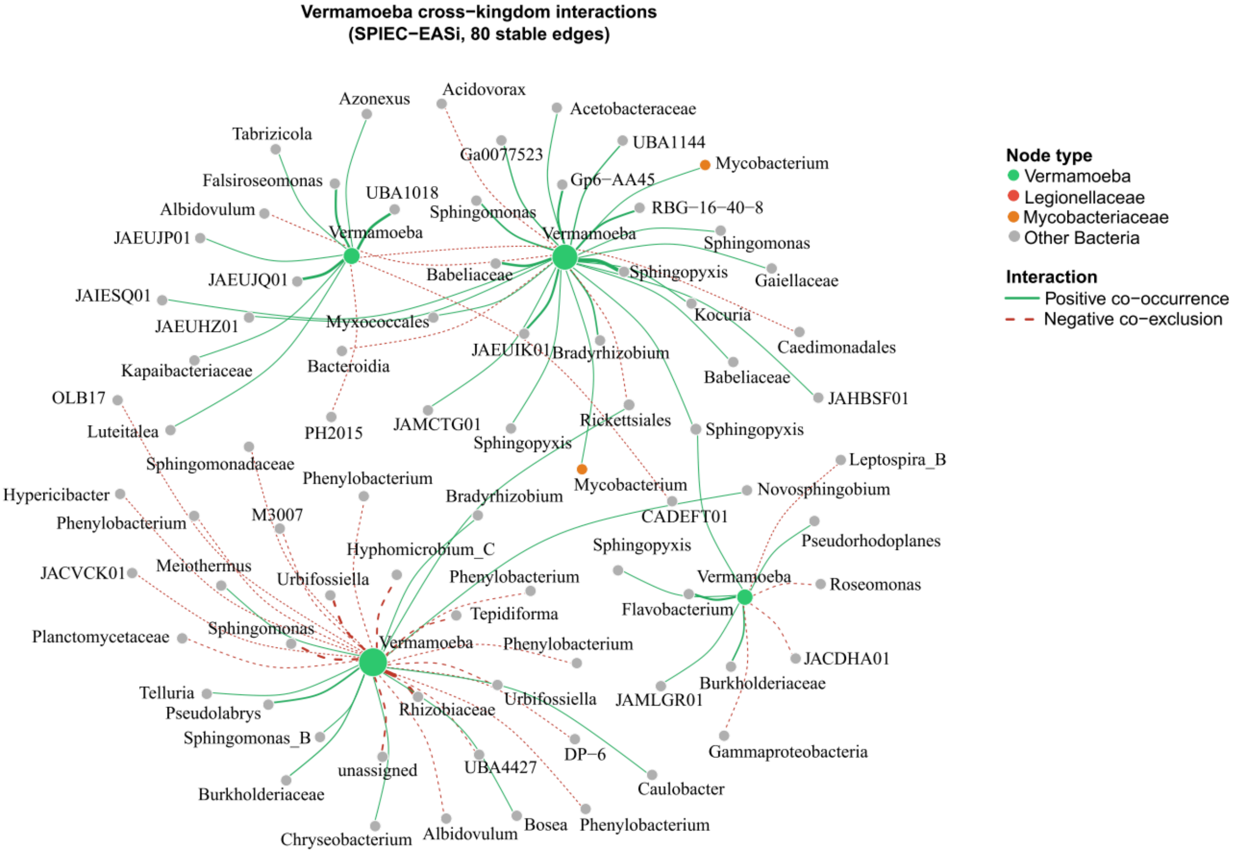


**Supplementary Figure 10:** Cross-kingdom co-occurrence network of Vermamoeba ASVs and their prokaryotic partners, inferred with SPIEC-EASI. Only stable edges identified by the StARS stability criterion are shown (n = 80). Node size is proportional to the number of stable edges. Legionellaceae are colored in red, Mycobacterium in orange and other bacteria in grey, while Vermamoeba in green. When taxonomic assignment was available, labels were at genus level, otherwise at the previous taxonomic level. Green edges indicate positive co-occurrence; red dashed edges indicate negative co-exclusion.
